# Between-Occupation Differences in Work-Related COVID-19 Mitigation Strategies over Time: Analysis of the Virus Watch Cohort in England and Wales

**DOI:** 10.1101/2022.10.31.22281732

**Authors:** Sarah Beale, Alexei Yavlinsky, Susan Hoskins, Vincent Nguyen, Thomas Byrne, Wing Lam Erica Fong, Jana Kovar, Martie Van Tongeren, Robert W Aldridge, Andrew Hayward, the Virus Watch Collaborative

## Abstract

**Background:** COVID-19 pandemic mitigations have had a profound impact on workplaces, however, multisectoral comparison of how work-related mitigations were applied across different phases of the pandemic are limited. This study aimed to investigate occupational differences in the usage of key work-related mitigations over time, and to investigate workers’ perceptions of these mitigations.

**Methods:** A survey covering the periods of late December 2020–February 2022 was developed and disseminated online to employed or self-employed participants in the Virus Watch study (*n*=6,279 respondents). Logistic regression was used to investigate occupation- and time-related differences in the usage of work-related mitigation methods. Responses regarding workers’ perceptions of mitigation methods were investigated descriptively using proportions.

**Findings:** Usage of work-related mitigation methods differed between occupations and over time, likely reflecting variation in job roles, workplace environments, legislation and guidance. Healthcare workers had the highest predicted probabilities for several mitigations, including frequent hand and surface hygiene (up to 0.61 [0.56, 0.66]), wearing face coverings (up to 0.80 [0.76, 0.84]), and employers providing face coverings for workers (0.96 [0.94, 0.98]) and other people on worksites (0.90 [0.87, 0.93]). There was a cross-occupational trend towards reduced mitigations during periods of less stringent national restrictions. The majority of workers across occupations (55-88%) agreed that most mitigations were reasonable and worthwhile; agreement was lower for physical distancing (39-44%).

**Interpretation:** While usage of work-related mitigations appeared to vary alongside stringency of national restrictions, agreement that most mitigations were reasonable and worthwhile remained substantial. Further investigation into the factors underlying between-occupational differences could assist pandemic planning and prevention of workplace COVID-19 transmission.

## Introduction

A diverse range of mitigation methods have been employed during the COVID-19 pandemic to reduce SARS-CoV-2 transmission in workplaces. These have included workplace closures, behavioural and environmental measures to reduce transmission (e.g., testing programmes and requirements to isolate from work if infectious, requirements or recommendations to wear face coverings, spatial reconfiguration to promote social distancing, ventilation), and promotion of COVID-19 vaccination by employers. Effective implementation of pandemic-related mitigation measures is likely to vary substantially by occupation due to variability in work environment, job roles and work cultures. Effective and proportional mitigations also vary by pandemic phase, and are likely influenced by time-varying legislation and guidance at the national, sectoral, and workplace levels. Occupational differences in SARS-CoV-2 infection risk have been observed across the pandemic (1-6), and continue to present concerns in terms of workforce disruption and long-term disability even with availability of safe and effective COVID-19 vaccines. Occupational differences in mitigation measures are likely to interact with workplace exposure to shape infection risk, and are consequently an important area for cross-sector investigation. Additionally, understanding how mitigation methods have been implemented across occupations is important to inform effective and economically viable planning for future public health threats. However, multi-occupation investigation into mitigation methods is currently limited.

Modelling, simulation and observational studies of workplace COVID-19 prevention and control strategies indicate that layered packages of mitigation methods - including gradual return to in-person working, asymptomatic testing, reduction of contact, and using personal protective equipment (PPE) – appear effective and more likely to reduce worker positivity compared to single measures (7–12). However, studies thus far have tended to focus on single workplaces, and many sectors outside of health and social care settings are underrepresented in the current literature (9). The effectiveness and feasibility of different packages of mitigations is likely to differ substantially across occupational sectors and roles, and multi-occupation observational studies are consequently warranted. In the UK – the regional focus of this study – this need is underscored by trade union reports indicating infrequent or inconsistent implementation of pandemic-related health and safety measures in a variety of workplaces during the first year of the pandemic (13,14). Empirical investigation into the implementation of work-related mitigations across a range of occupational groups would both provide potential insight into occupational differences in infection risk and possible areas for intervention, and provide evidence to plan for future public health emergencies.

This study aimed to investigate between-occupation and time-related differences in the implementation of workplace mitigation methods in England and Wales during key phases of pandemic-related national legislation between late December 2020 (third national lockdown in both nations) and late February 2022 (after relaxation of most pandemic restrictions). Specific objectives were:

1. To investigate how implementation and uptake of key work-related mitigations varied by occupation and, where relevant, by pandemic phase.
2. To investigate which COVID-19 mitigations methods were perceived as reasonable and worthwhile by workers in different occupations during the third national lockdown (late December 2020 – March 2021) and during a period of relaxed restrictions in February 2022.

## Methods

### Ethics Approval

The Virus Watch study was approved by the Hampstead NHS Health Research Authority Ethics Committee: 20/HRA/2320, and conformed to the ethical standards set out in the Declaration of Helsinki. All participants provided informed consent for all aspects of the study.

### Participants

Participants were an adult sub-cohort of the Virus Watch longitudinal cohort study(15). The Virus Watch study recruited households who met the following inclusion criteria using social media, SMS, and personalised postal recruitment supported by general practices: ordinarily resident in England or Wales, household between 1-6 people (due to limitations on survey infrastructure), had access to the internet and to an email address, and at least one household member able to complete surveys in English. Further details of the main Virus Watch cohort and recruitment are provided in the study protocol(15). Participants from the main cohort were included in the present study if they met the following further criteria: an adult ≥16 years, who responded to a survey sent on 22 February 2022 regarding mitigation methods in the workplace, who was employed or self-employed and not on full-time furlough during at least part of the survey period, and who reported a valid, consistent occupation through the survey period (when employed or self-employed).

Participants who reported changing occupations between survey periods on the questionnaire (*n*=381) were excluded using skip logic due to limitations with the survey infrastructure and complexity (see ‘Outcomes’ section below for further description of survey).

### Exposure

Participants’ provided their main occupation as free text during registration with the Virus Watch cohort and at the beginning of the survey underlying this study (sent in February 2022). We used responses to the February 2022 survey as a preferred source due to direct coverage of the survey period. Responses from the baseline survey were used where participants reported being employed or self-employed but did not provide a classifiable occupation (*n*=586). As the survey was displayed only to participants who indicated a consistent occupation throughout the survey period, we assumed that the baseline survey was likely to be representative and included these participants to strengthen sample size.

To classify occupation, we assigned UK Standard Occupational Classification (SOC) 2020(16) codes using semi-automatic processing in Cascot Version 5.6.3. If participants reported multiple occupations, the first listed occupation was used. We then used SOC codes to classify participants into the following occupational groups, which aimed to reflect workplace environment which retaining the overall structure of SOC-defined skill groupings where possible: administrative and secretarial occupations; healthcare occupations; indoor trade, process & plant occupations; leisure and personal service occupations; managers, directors, and senior officials; outdoor trade occupations; sales and customer service occupations; social care and community protective services; teaching, education and childcare occupations; transport and mobile machine operatives; and other professional and associate occupations.

The most prevalent SOC-2020-defined occupations for participants in this study are reported in Supplementary Table 1. Analyses could not be further disaggregated by specific occupations due to sample size limitations.

### Outcomes

All outcomes were derived from responses to a one-off survey sent on 22 February 2022 and displayed to all cohort participants over 16 years of age. Questions aimed to cover key aspects of work-related transmission risk and associated mitigations based on contemporary understanding of transmission pathways of SARS-CoV-2 (17–19) and UK governmental sources regarding COVID-19 legislation and recommendations applicable to workplaces(20,21). The full survey is given in the Supplementary Materials (‘Virus Watch Work-Related Mitigations Survey (February 2022)’).

The first section of the survey comprised items regarding implementation and usage of key COVID-related mitigation methods in the workplace. Items addressed key mitigation methods including social distancing, ventilation, usage of face coverings, usage of LFTs, surface and hand hygiene, and promotion of COVID-19 vaccination. Items applicable across multiple phases of the pandemic and liable to substantial change were asked separately for the following periods, reflecting broad phases of restrictions: late December 2020 – March 2021 (third national lockdown in England and Wales), July – December 2021 (most restrictions relaxed during this period in both nations), late December 2021 – January 2022 (Omicron/Phase 2 restrictions in both nations), or current survey period (most restrictions relaxed in both nations). The survey was limited to the period between late December 2020 to February 2022 to balance recall bias with collecting information across key periods of national legislation. Some items– particularly those relevant to risk-related workplace features – were adapted from previous sources including the COVID-19 Job Exposure Matrix (22)^1^, other Virus Watch surveys, and items about the Flu Watch prospective cohort study (23); permission to use or adapt items was sought where required. Supplementary Table 2 reports the source that each item was adapted from. Skip logic was used to display only relevant periods when the participant reports being employed or self-employed and to display items relating to the workplace environment only for periods with in-person attendance; skip logic and the consequent questionnaire structure is detailed further in Supplementary Figure 1.

In the second section of the survey, participants rated how reasonable and worthwhile they believed key mitigation methods to be in their workplace during the third national lockdown (most stringent period of restrictions covered by the survey) and the current phase of the pandemic at the time of the survey (late February 2022 after relaxation of most pandemic-related restrictions). Items were rated on a five-point Likert-type scale: Strongly disagree (not at all reasonable or worthwhile) – Strongly agree (very reasonable and worthwhile), with the additional potential response ‘Not possible/relevant in my job’.

### Covariates

Where required (see Statistical Analyses), models were adjusted for the following covariates: age (<30, 30-39, 40-49, 50-59, 60+ years), sex at birth, employment status (working up to 20 hours per week, working 20-35 hours per week, working more than 35 hours per week), and clinical vulnerability status (vulnerable versus non-vulnerable, based on reporting of any medical condition classified by Public Health England/UK Health Security Agency, the Department of Health and Social Care, and the Joint Committee on Vaccination and Immunisation (27,28) to denote vulnerability to severe COVID-19. Employment status was entered as a time-varying covariate. Age and sex were derived from responses to the Virus Watch registration survey, employment status was drawn from the February 2022 survey and clinical vulnerability was derived based on data sources detailed elsewhere(24).

### Statistical Analyses

Ordinal or binomial logistic regression was used to investigate between-occupational differences for all outcomes in the first section of the survey. For items measured across multiple time periods, cluster-robust standard errors were used to account for within-individual clustering. Wald tests based on a cluster-robust estimate of the variance matrix were used to assess evidence of an interaction between occupational group and time.

Based on the VanderWeele principle of confounder selection(25) and adjustment sets for previous analyses of workplace attendance during the pandemic(26), the model for in-person workplace attendance was adjusted for age, sex, clinical vulnerability, and employment status. This model was not adjusted for vaccination status, as participants’ vaccination status was not assumed to alter general patterns of attendance across the broad time-periods represented in the survey and there was limited variation in vaccination status during the survey period. The effect of socio-demographic factors on other outcomes was presumed to occur via the impact of occupation, time and/or workplace attendance, and subsequent workplace-related items were only displayed to participants who attended in-person during a given time period due to the nested survey structure (Supplementary Figure 1). ‘Unsure’ responses were dropped from relevant regression models to retain ordinal scales for most items. Complete case analysis was performed based on available responses for each question and missing data were limited for covariates (see Table 1); the number of respondents varied by question due to the nested structure of the items (Supplementary Figure 1). The number of total respondents and ‘Unsure’ (excluded) responses per item are reported in Supplementary Table 3.

**Table 1.** Demographic Characteristics of Participants

| Characteristic | Participants (Current Study)<br>N = 6,279<sup>1</sup> | Workers in Virus Watch Cohort<br>N = 20,258<sup>1</sup> |
| --- | --- | --- |
| **Occupation** |  |  |
| Administrative & Secretarial | 880 (14%) | 2,539 (13%) |
| Healthcare | 584 (9.3%) | 1,686 (8.3%) |
| Indoor Trades, Process & Plant | 411 (6.5%) | 1,405 (6.9%) |
| Leisure & Personal Service | 304 (4.8%) | 1,014 (5.0%) |
| Managers, Directors & Senior Officials | 509 (8.1%) | 1,653 (8.2%) |
| Other Professional & Associate | 1935 (31%) | 6,539 (32%) |
| Outdoor Trades | 193 (3.1%) | 471 (2.3%) |
| Sales & Customer Service | 295 (4.7%) | 1,058 (5.2%) |
| Social Care & Community Protective Services | 368 (5.9%) | 1,117 (5.5%) |
| Teaching, Education & Childcare | 611 (11%) | 2,297 (11%) |
| Transport & Mobile Machine | 139 (2.2%) | 479 (2.4%) |
| **Age** |  |  |
| <30 | 303 (4.8%) | 1,796 (8.9%) |
| 30-39 | 535 (8.5%) | 3,457 (17%) |
| 40-49 | 1,112 (18%) | 4,402 (22%) |
| 50-59 | 2,164 (34%) | 5,728 (28%) |
| 60+ | 2,165 (34%) | 4,875 (24%) |
| **Sex** |  |  |
| Female | 3,658 (58%) | 11,299 (55.8%) |
| Male | 2,603 (41%) | 8,923 (44%) |
| Unknown/Other | 18 (0.3%) | 36 (0.2%) |
| **Clinically Vulnerability** |  |  |
| Clinically vulnerable | 2,279 (36%) | 7,031 (35%) |
| Not clinically vulnerable | 4,000 (64%) | 13,227 (65%) |
| **Ethnicity** |  |  |
| White British | 5,401 (88%) | 16,411 (81%) |
| White Other | 466 (7.6%) | 1,855 (9.3%) |
| Mixed | 71 (1.2%) | 346 (1.7%) |
| South Asian | 86 (1.4%) | 906 (4.5%) |
| Other Asian | 46 (0.8%) | 203 (1.0%) |
| Black | 38 (0.6%) | 214 (1.1%) |
| Other Ethnicity | 25 (0.4%) | 118 (0.6%) |
| Unknown | 146 (2.3%) | 205 (1%) |
| **IMD Quintile** |  |  |
| 1 | 546 (8.8%) | 2,081 (10%) |
| 2 | 973 (16%) | 3,480 (17%) |
| 3 | 1,266 (20%) | 4,089 (20%) |
| 4 | 1,611 (26%) | 4,944 (24%) |
| 5 | 1,800 (29%) | 5,390 (27%) |
| Unknown | 83 (1.3%) | 274 (1%) |
| **Region** |  |  |
| East Midlands | 600 (9.6%) | 1,799 (8.9%) |
| East of England | 1,222 (19%) | 3,889 (19%) |
| London | 860 (14%) | 3,565 (17%) |
| North East | 254 (4.0%) | 882 (4.4%) |
| North West | 617 (9.8%) | 2,031 (10%) |
| South East | 1,261 (20%) | 3,846 (19%) |
| South West | 517 (8.2%) | 1,388 (6.9%) |
| Wales | 199 (3.2%) | 547 (2.7%) |
| West Midlands | 354 (5.6%) | 1,058 (5.2%) |
| Yorkshire and The Humber | 312 (5.0%) | 979 (4.8%) |
| Unknown | 83 (1.3%) | 274 (1%) |

<sup>†</sup>n (%)

For the survey items pertaining to participants’ perceptions of mitigation methods in the workplace, we calculated response proportions stratified by occupational group and time period. Inferential analyses were not performed for these items as they were intended to illustrate how workers’ perceptions varied across mitigation methods with a view to provide evidence to inform future interventions. Direct occupational comparison was not the objective of this analysis regarding workers’ perceptions. This is in contrast to the first section of the survey, which was intended to investigate occupational differences in potentially risk-relevant features and mitigation methods over time.

## Results

A total of 6,279 participants were included in the study. Participants’ demographic features are reported in Table 1, along with all workers with known occupation in the Virus Watch cohort. Demographic features were similar between survey participants and the full cohort of workers, with some increased representation of older workers and those of a White British background amongst survey respondents. Participant selection illustrated in Supplementary Figure 2 and employment status over time in Supplementary Table 4.

Based on Wald tests for the inclusion of an interaction term between occupational group and time, an interaction term was included in the final model for all outcomes pertaining to multiple time periods (Wald *p*<0.001) excluding frequency of hand hygiene (*p*=0.09) and surface hygiene (*p*=0.17) and degree of precautions taken during breaks (*p*=0.30), which demonstrated main effects for occupation and time. Where identified, these interactions indicated that the frequency of the outcomes changed over time differentially by occupation.

### Workplace Sharing and Social Distancing

The average number of in-person workdays changed differentially over time between occupations (Supplementary Figure 3). Across all periods, probability of full-time in-person workplace attendance was highest for tradespeople, transport and mobile machine operatives and leisure and personal service workers (range for these groups across periods: predicted probability (PP) range 0.43 [0.37, 0.50] – 0.58 [0.52, 0.64]) and lowest for Other Professional and Associate occupations (PP range across periods: 0.05 [0.05,0.06] – 0.14 [0.13,0.16]). Teaching, education and childcare workers exhibited substantial change over time, with in-person attendance 5+ days per week becoming common after the third national lockdown. Findings were similar in the unadjusted model (Supplementary Figure 4).

Intensity of workspace sharing varied between occupations over time and was most intense for teaching, education, and childcare occupations and sales and customer service occupations; however, space sharing was common across occupations (Supplementary Figure 5). Predicted probabilities for the workspace always being socially distanced were relatively low across all occupations (Supplementary Figure 6), even during the third national lockdown: PP range for this period 0.05 (0.04, 0.07) to 0.22 (0.17, 0.26). Healthcare workers and teaching, education and childcare workers persistently demonstrated the highest probabilities of reporting no social distancing at work (PP range 0.15 [0.12, 0.17] to 0.24 [0.21, 0.27]), with confidence intervals exceeding estimates for most other groups.

Strategies used in the workplace to promote social distancing also varied by occupation (Figure 1). All occupations had predicted probabilities around or above 50% for reconfiguring the workspace (PP range 0.60 [0.46, 0.74] to 0.86 [0.81, 0.90]), limiting occupancy (PP range 0.65 [0.59,0.71] to 0.75 [0.81 to 0.89]), using one-way systems (PP range 0.49 [0.35, 0.63] to 0.75 [0.70,0.79]), and using posters/reminders (PP range 0.48 [0.34, 0.62 to 0.89 [0.84, 0.94]). Use of screens/barriers was more likely in sales and customer service occupations (PP 0.82 [0.75, 0.88]) than for any other group. Tradespeople tended to have lower probabilities than many other groups across a range of social distancing methods. Teaching and childcare workers had the highest probability of reporting staggered breaks (PP 0.60 [0.55, 0.65]) and use of workplace bubbles (PP 0.64 [0.59, 0.68]) and the lowest probability of reporting staggered shifts (PP 0.12 [0.08, 0.15]); confidence intervals indicated considerable between-occupational overlap for other groups.

**Figure 1.**
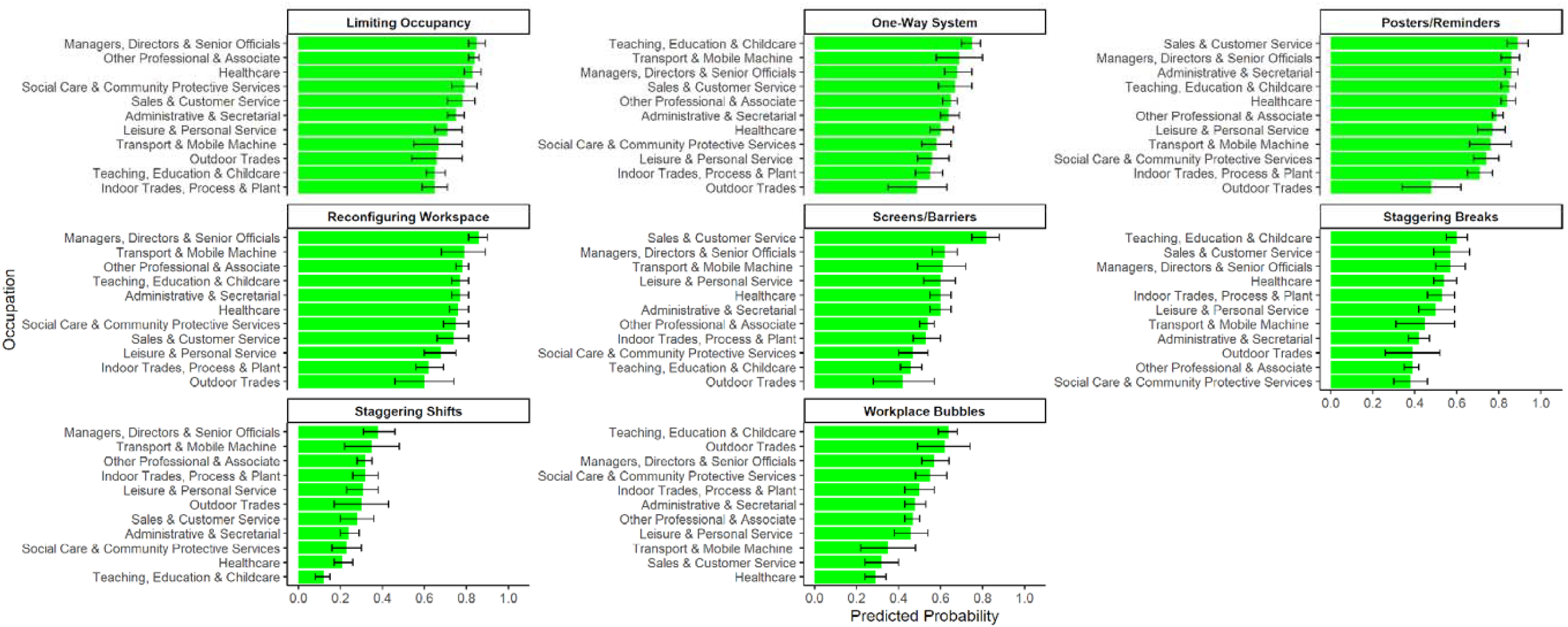
Strategies to Reduce Contact-Related Risk in the Workplace: Predicted probabilities by occupation

### Ventilation

Working environment by occupation is reported in Supplementary Figure 6. Except for outdoor tradespeople and transport and mobile machine operatives, the majority of participants across occupations reported working primarily indoors. Amongst participants who worked at least partly indoors, physical ventilation was the most commonly-reported method across groups (Supplementary Figure 7): PP range 0.64 (0.56, 0.73) to 0.93 (0.85, 1.00); predicted probabilities were higher for teaching, education and childcare and transport and mobile machine operatives than in most other groups. Predicted probabilities for mechanical ventilation (Supplementary Figure 7) ranged from 0.25 (0.21, 0.29) to 0.60 (0.56, 0.63), and for air purifiers or filters ranged from 0.12 (0.05, 0.15) to 0.28 (0.22, 0.34). Teaching, education and childcare workers had the lowest likelihood of reporting these measures and managerial and Other Professional and Associate occupations the highest.

### Hand and Surface Hygiene

Frequency of touching shared surfaces and objects is reported in Supplementary Figure 8. The probability of very frequently touching shared surfaces and objects was lower in outdoor trades (0.09 [0.06, 0.12]) and higher in healthcare, sales and customer service, leisure and personal service, and teaching, education and childcare occupations (PP range 0.39 [0.35, 0.44] – 0.46 [0.41, 0.50]) compared to all other groups.

Frequency of hand hygiene in the workplace varied substantially by occupation (Figure 2a) and over time (Figure 2b) with no interaction. Healthcare occupations tended to have greater hand hygiene frequency, with a higher probability of reporting washing or disinfecting their hands >10 times per workday than any other group (PP 0.61 [0.56, 0.66]). Outdoor trades (PP 0.61 [0.52, 0.70]) and Other Professional and Associate occupations (PP 0.57 [0.54, 0.60]) had higher probability of infrequent hand hygiene (0-5 times per workday) than any other group. Confidence intervals indicated considerable overlap in hand hygiene frequency for other occupations. Across all occupations, the probability of reporting infrequent hand hygiene increased between the third national lockdown (PP 0.36 [0.43, 0.37]) and subsequent periods (PP range 0.42 [0.40, 0.44] – 0.43 [0.41, 0.45]), and increased substantially again in February 2022 (0.48 [0.46, 0.50]); the probability of reporting very frequent hand hygiene decreased correspondingly over time.

**Figure 2.**
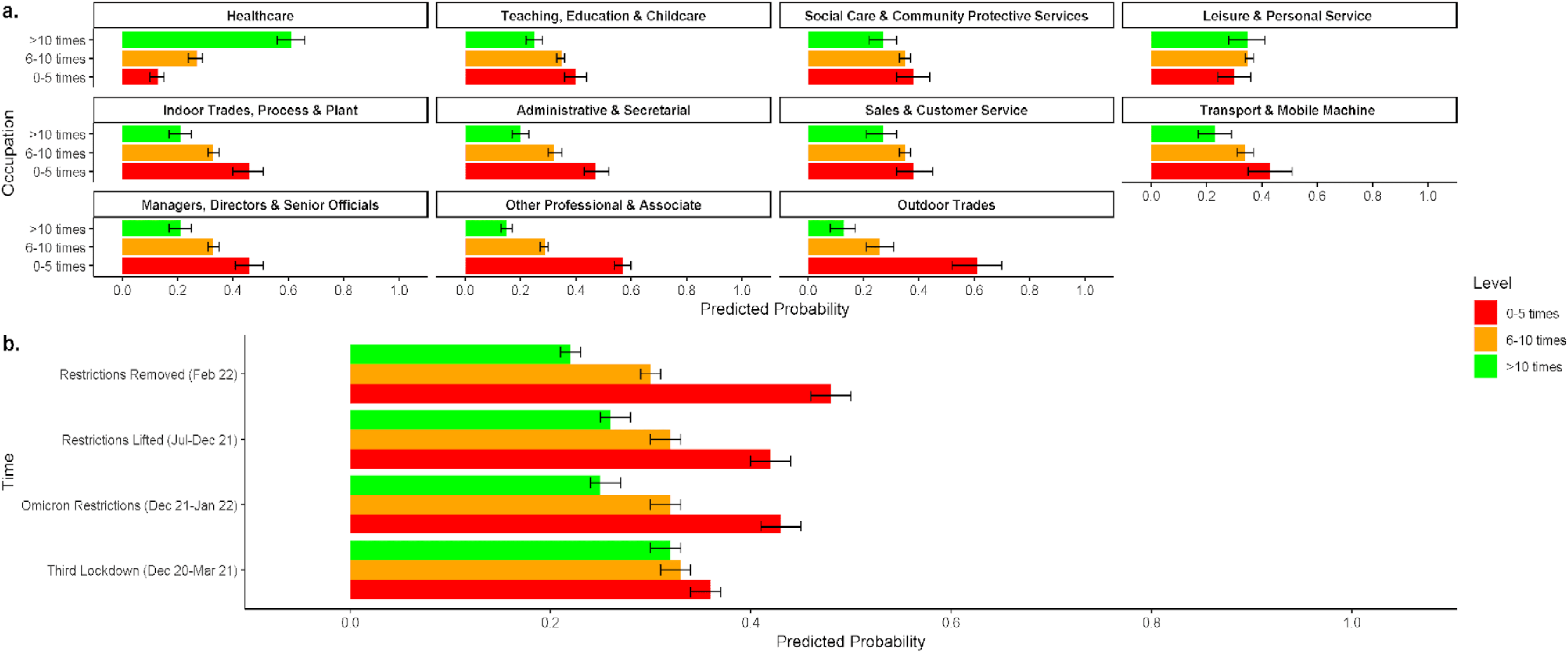
Frequency of Handwashing at Work: Predicted probabilities by occupation (a) and over time (b)

Similar occupational and time-based patterns were observed for frequency of surface hygiene (Supplementary Figures 9a and 9b). Frequency of surface hygiene decreased across survey periods (PP range for never disinfecting: 0.22 [0.20, 0.24] during third national lockdown to 0.30 [0.28, 0.31] in late February 2022).

### Face Coverings

Workers in healthcare, teaching, education and childcare, social care and community protective service, leisure and personal service, and sales and customer service occupations were more likely to self-report always using a face covering at work (Supplementary Figure 10) compared to other groups between the third national lockdown and the Omicron restriction period (PP range 0.41 [0.35, 0.47] to 0.80 [0.76, 0.84]). The probability of always using a face covering decreased for these groups by February 2022 (range 0.24 [0.17, 0.32] – 0.31 [0.25, 0.37]), excluding healthcare workers for whom it remained high (0.71 [0.66, 0.75]). Healthcare workers also had greater probability of reporting that other people on the worksite always wore face coverings compared to any other occupational group (Supplementary Figure 11). However, following cross-occupational trends, this probability decreased within this group in February 2022 (0.48 [0.43, 0.52]) compared to the initial three survey periods (range 0.63 [0.57, 0.67] – 0.68 [0.64, 0.73]).

Workplaces were more likely to provide face coverings to healthcare workers (PP 0.96 [0.94, 0.98]) and other people attending healthcare settings (PP 0.90 [0.87, 0.93]) than any other occupational group during the survey period (Supplementary Figure 12). Outdoor trade workers reported that their workplaces were less likely to provide face coverings to workers (PP 0.55 [0.43, 0.67]) or other people attending the worksite (PP 0.44 [0.31, 0.57]) compared to most other occupations.

### Breaks and Work-Related Social Activities

Typical contact with others during breaks is reported in Supplementary Figure 13. Spending breaks indoors with other people was relatively common across occupations (PP range 0.21 [0.13, 0.29]-0.53 [0.48, 0.57]). Workers commonly reported that fewer pandemic-related precautions were taken during breaks (Figure 3) compared to active work across occupations: PP range 0.39 (0.35, 0.44) to 0.54 (0.46, 0.61). For all groups, the probability of reporting fewer precautions during breaks increased over time, ranging from 0.30 (0.28, 0.32) during the third national lockdown to 0.59 (0.57, 0.61) in late February 2022. There was no interaction between occupation and time for this outcome.

**Figure 3.**
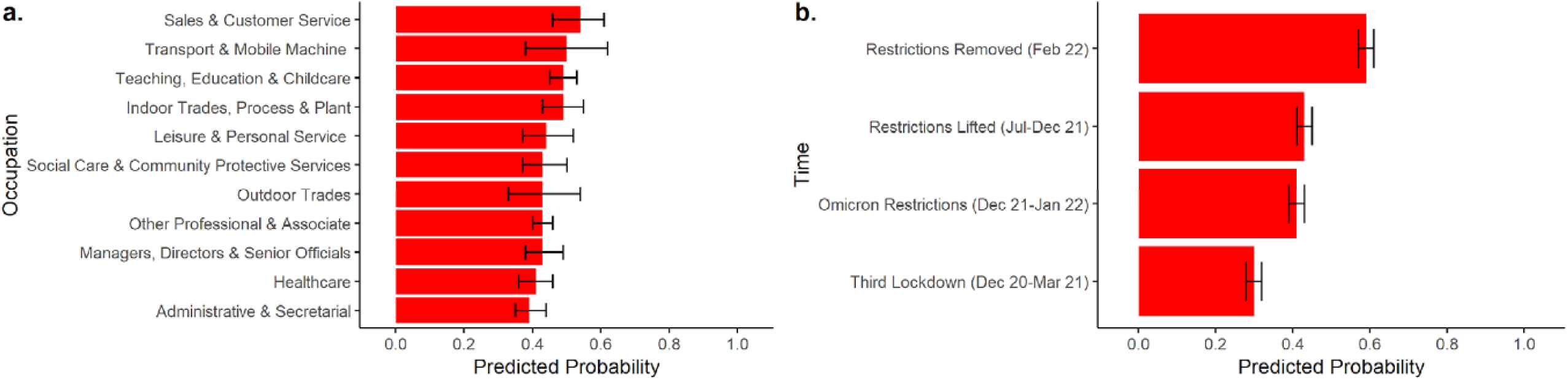
Fewer Precautions during Breaks Compared to Active Work?: Predicted probabilities by occupation (a) and time (b)

Probabilities of work-related social gatherings (“social events outside of working hours organised by the workplace, or social events on work premises including food and/or drinks”) differed between occupations across survey periods (Supplementary Figure 14). The most common response across all occupational groups and time periods was that social gatherings never occurred: PP range 0.48 (0.42, 0.55) – 1.00 (1.00, 1.00). Confidence intervals indicated an increasing tendency towards reporting social gatherings over time for most groups except tradespeople, sales and customer service occupations, and transport and mobile machine operatives.

### Lateral Flow Testing

Occupational groups differed over time in their probability of regular LFTs being required or recommended to attend work (Figure 4). Teaching, education and childcare workers had the highest probability of requiring an LFT to attend the worksite across survey periods (range 0.45 [0.41, 0.48)]–0.54 [0.50, 0.59]). Tradespeople, transport and mobile machine operatives, and sales and customer service occupations had the highest probabilities of reporting no explicitly-discussed workplace testing strategy (range for these groups across periods 0.51 [0.38,0.65] – 0.64 [0.52,0.77]).

**Figure 4.**
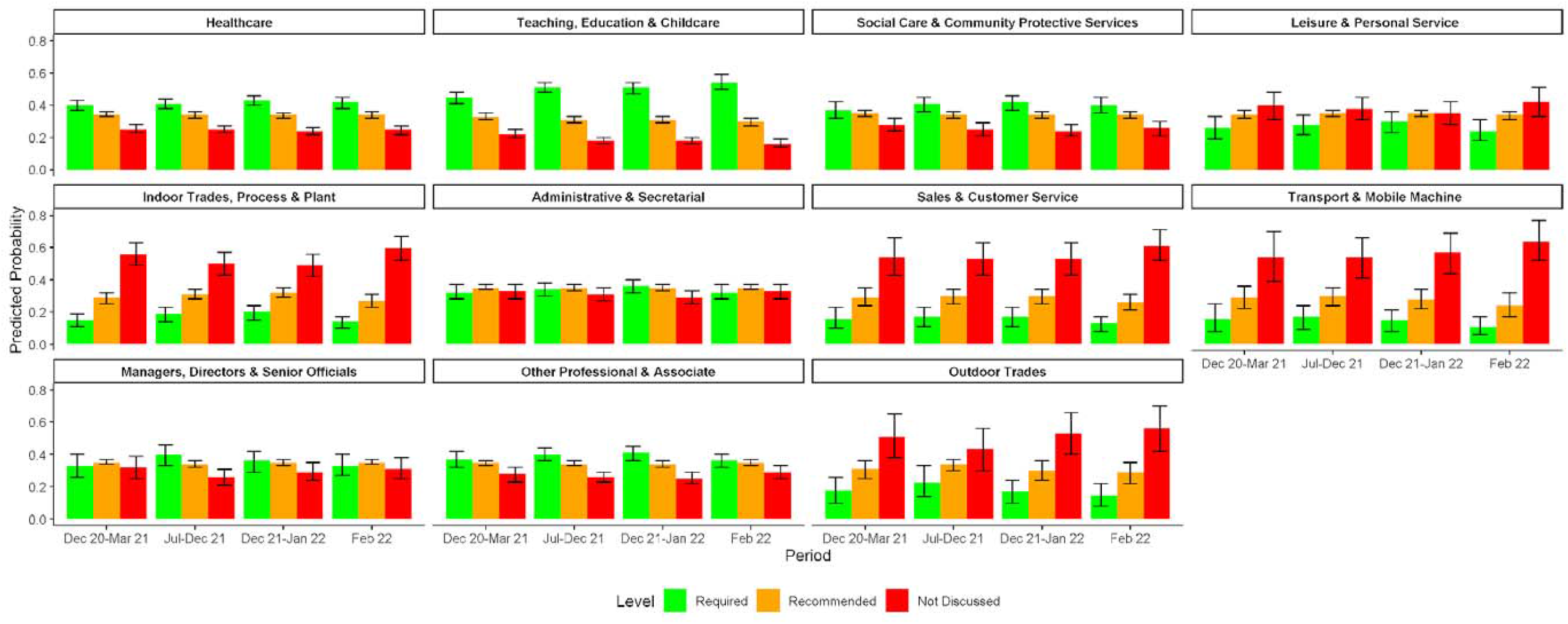
Regular Lateral Flow Testing for Workplace Attendance: Predicted probabilities by occupation over time

Workplace provision of LFTs at any point during the study period (Supplementary Figure 15) ranged from 0.08 (0.02, 0.14) to 0.48 (0.44, 0.53) for on-site testing, and from 0.23 (0.15, 0.31) to 0.83 (0.80, 0.87) for at-home test kits. Occupational patterns for LFT provision were similar to those described above regarding testing strategy.

### Workplace Promotion of COVID-19 Vaccination

Workplace strategies to promote COVID-19 vaccination varied between occupations (Figure 5). The most common method of promoting vaccination overall was providing time off work to attend vaccination appointments (PP range 0.50 [0.38, 0.62] – 0.86 [0.82, 0.89]), followed by use of promotional materials in the workplace (PP range 0.22 [0.13, 0.30] to 0.78 [0.74, 0.82]), and mandatory vaccination policies (PP range 0.03 [0.00, 0.06] to (0.49 [0.44, 0.54]). Provision of vouchers was rare across all groups (PP range 0.00 [0.00, 0.00] – 0.06 [0.03, 0.08]). Across strategies, healthcare and social care and community protective service workers tended to have higher probabilities – with marked differences compared to all other occupational groups for mandatory vaccination and use of promotional materials. Tradespeople and transport and mobile machine operatives demonstrated relatively low probabilities across strategies. Probability of reporting other strategies to promote vaccination outside of those explicitly included in the questionnaire was relatively low across occupations (0.03 [0.01, 0.08] – 0.18 [0.13, 0.23]), with the highest estimate in healthcare workers. Specific qualitative information about other strategies was not available based on the questionnaire.

**Figure 5.**
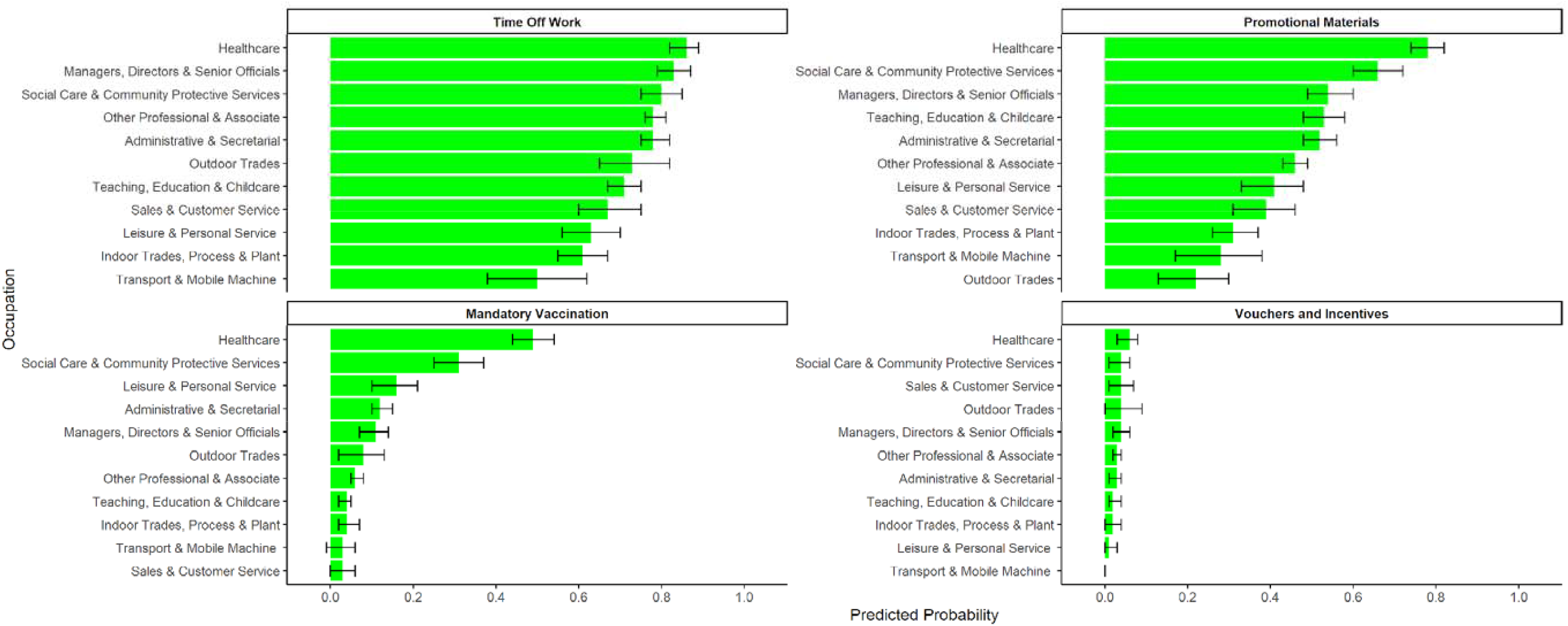
Strategies to Promote COVID-19 Vaccination: Predicted probabilities by occupation

### Perception of Work-Related Mitigation Methods

Workers’ perceptions of key work-related mitigation methods are reported by occupation in Figure 6 for the third national lockdown and Figure 7 for late February 2022. Across all occupations at both timepoints, ≥50% of participants agreed or strongly agreed with each measure except physical distancing. Patterns of agreement were similar across occupations, and respondents agreed or strongly agreed with the following measures in descending order: regular testing (88% during third national lockdown and 84% in February 2022), requiring face coverings for workers (88% and 84%), proof of vaccination for workers (86% and 74%), ventilation (83% and 62%), requiring face coverings for non-workers attending the worksite (83% and 62%), screens/barriers (79% and 60%), working from home (76% and 58%), surface cleaning (68% and 54%), proof of vaccination for non-workers attending the worksite (e.g., customers, clients, patients) (62% and 55%), and physical distancing (44% and 39%).

**Figure 6.**
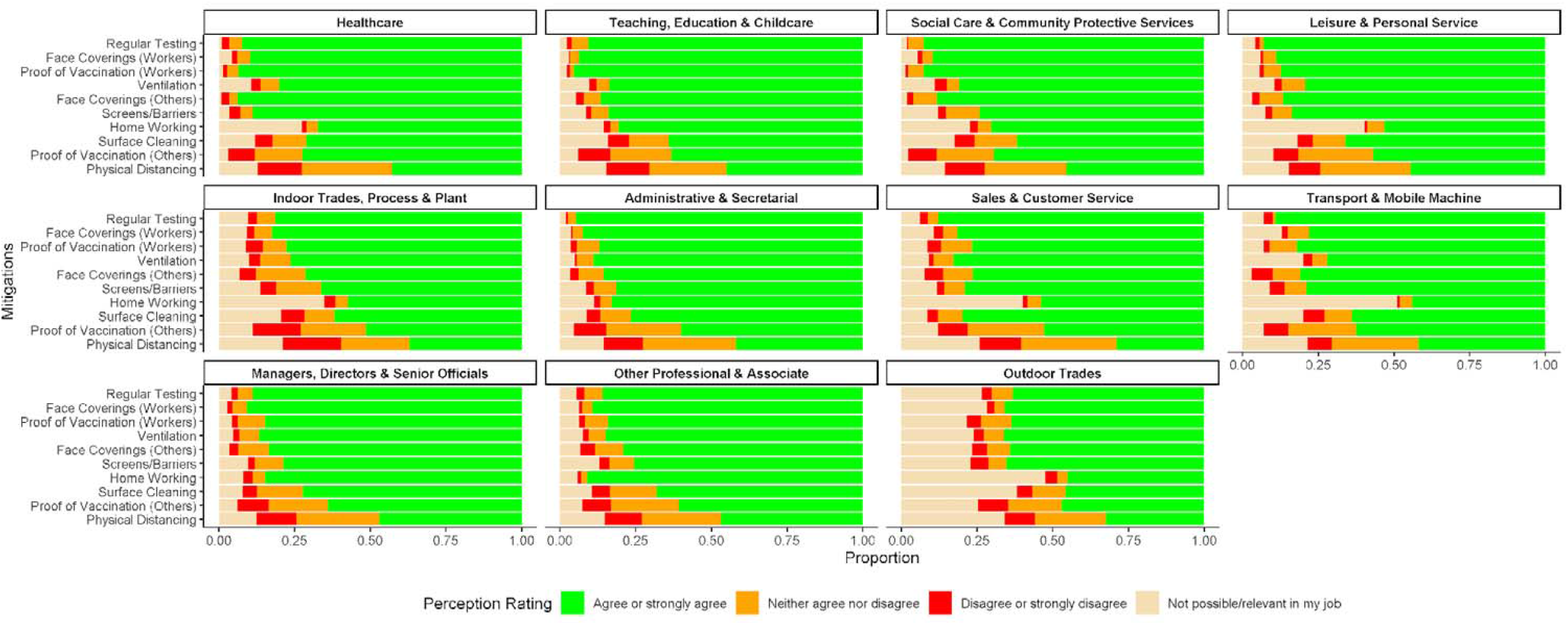
Perceptions of Work-Related Mitigation Methods during the Third National Lockdown: Proportions by occupation

**Figure 7.**
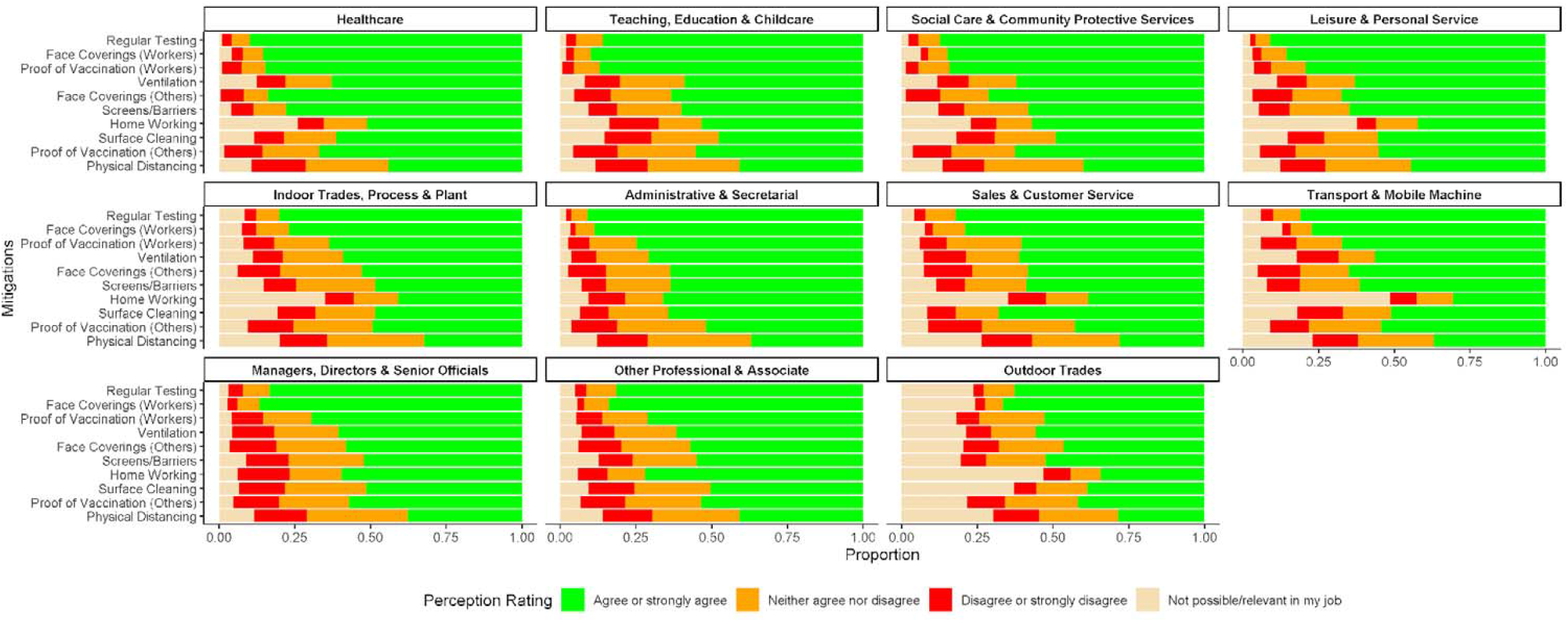
Perceptions of Work-Related Mitigation Methods in late February 2022: Proportions by occupation

Home working had the highest proportion of ‘not relevant or possible’ responses across all respondents (19% for third national lockdown and 18% versus late February 2022, Appendix 31), particularly for tradespeople, transport and mobile machine operatives, leisure and personal service, and sales and customer service workers at both timepoints. Outdoor tradespeople had relatively high levels of reporting that measures were not relevant or possible for their workplace across measures at both timepoints (range across mitigations: 18%-48%).

## Discussion

### Key Findings and Interpretation

This study found substantial between-occupational differences in risk-relevant workplace features and related mitigations, with patterns of variation corresponding to both occupational features and national legislation. In line with previous findings regarding earlier phases of the pandemic(26), there was a cross-occupational trend towards more intense space sharing and fewer mitigations during periods of less intense national restrictions despite high levels of community transmission.

The survey period corresponded with the third and fourth pandemic waves in England and Wales. Longitudinal studies of occupational infection risk found fewer between-occupational differences in risk during these waves than earlier in the pandemic, which were primarily attributed to higher levels of population mixing across venues reducing the specific importance of the workplace (5,6). However, teaching occupations – who had evidence of persistently elevated risk across the survey period (5,6) – also demonstrated high levels of potential workplace risk and relatively low implementation of some key mitigations (i.e., ventilation) in the present study. Wide-ranging mitigations in some high-risk occupational groups (e.g., healthcare) may have mitigated workplace infection risk effectively. Direct investigation of the relationship between mitigations and infection risk was beyond the scope of this study.

Furthermore, preventing infection risk in the workplace across sectors continues to be relevant due to the ongoing risk of long-term post-infection sequelae, disruption due to workplace absences, and potential future public health threats. Consequently, understanding occupational and temporal change in how specific measures were implemented and viewed by workers is warranted, and specific findings are reviewed below.

#### Workplace Behavioural and Environmental Mitigations

In-person workplace attendance appeared to correspond with previous classifications of ability to work from home(27) and – for some occupations – appeared to reflect sectoral re-openings; this corroborates findings from earlier phases of the pandemic(26). The relevance of work-related mitigations was illustrated by potential transmission risk, as workplaces tended to be shared and social distancing was often inconsistent, even during periods of stringent national restrictions.

Occupational differences in the implementation of behavioural and environmental mitigations appeared to reflect differences in job roles, working environment, and related legislation and guidance. For example, social distancing was least frequently reported in teaching, education and childcare and healthcare occupations, where intense contact with others may be unavoidable. Public-facing workers such as those in healthcare and teaching, education and childcare occupations correspondingly reported the highest frequency of using face coverings at work. Most occupations – excluding healthcare – reported a trend towards decreased usage of face coverings over time.

Persistent usage and workplace provision of face coverings to workers and other people in healthcare settings likely reflects ongoing regulations aimed at infection prevention in healthcare environments(28). Due to burden-related limitations, further detail about types and contexts in which face coverings were used was beyond the scope of this survey.

Frequency of hand and surface hygiene also appeared to reflect similar occupational differences and pre- and peri-pandemic infection control protocols (28,29), with the highest frequencies in groups who reported touching shared surfaces and objects often, e.g., healthcare and leisure and personal service workers. Hand and surface hygiene decreased over time for all occupational groups, which may also impact transmission of other infections in the workplaces spread via this route.

Across occupations, workers tended to report that mitigations such as social distancing tended to reduce over time. This cross-occupational trend may reflect decreased stringency of national restrictions, though this cannot be directly inferred from the current study. A previous investigation into workplace contact patterns during earlier pandemic periods in the UK found that contact behaviour appeared to mirror the stringency of restrictions(26). Workers also commonly reported that people in their workplace took fewer precautions during breaks compared to active work, with this trend becoming more prominent over time compared to the third national lockdown. Effective support for maintaining protective measures during breaks is a relevant area for further inquiry to inform pandemic and outbreak management planning. Workplace social gatherings, where social distancing may be more difficult to maintain, also increased across time for most occupational groups but were relatively uncommon across occupations, potentially indicating that this may not be a major factor underlying between-occupational differences in transmission. Direct investigation of this relationship was not possible due to data-related limitations.

Ventilation is an important environmental mitigation to prevent transmission of respiratory viruses with minimal reliance on individual behaviour. In this study, physical ventilation was commonly reported (predicted probabilities of 64-93% across occupations). Mechanical ventilation and air filtration were less common, with the highest usage in managerial and other professional occupations and notably low usage in teaching, education and childcare occupations. While the effectiveness of ventilation depends on properties of the workplace environment, mechanical ventilation tends to be more effective than physical methods(30). Given the high-intensity contact associated with teaching and childcare occupations, proactively scaling-up ventilation in schools may be a beneficial public health measure to reduce transmission of respiratory viruses and improve sectoral resilience in the face of future public health threats.

#### Lateral Flow Testing

Lateral flow testing was a flagship element of the national COVID-19 response in England and Wales (31,32) aimed at identifying infectious individuals and preventing them from mixing with others. High costs were accrued by government-subsidised testing programmes (33,34). In this survey, testing was most commonly required and provided for healthcare, social care and community protective service, and teaching, education and childcare professionals. This likely reflected national proactive test availability, guidance and communications targeting these sectors, for whom testing was made available earlier than for the general population.

Despite free availability for all individuals and businesses from April 2021, workers in non-target occupational groups commonly reported no explicit guidance around testing (predicted probability range across periods 25%-64%) and low workplace provision of on-site (8-34%) or at-home testing for workers (29-45%). This was particularly likely for tradespeople and transport and mobile machine operatives, despite persistently high in-person workplace attendance. These findings indicate occupational inequalities that add to emerging evidence around sociodemographic inequalities in the implementation and uptake of mass testing programmes(35). Clear communication and support around testing at the employer and government level is likely to be crucial for future mass testing programmes, as unclear recommendations and fears around income loss or disruption to work may reduce workers’ engagement with testing(35,36). Workers’ personal usage of LFT devices was not investigated in this survey, but other behavioural surveys suggest low general population usage of LFTs in the UK (37). Further investigation into why workplace-level guidance and test provision remained low across many sectors is recommended in order to identify factors at the policy level, employer level, and worker level that may affect the implementation of these programmes.

#### Workplace Promotion of Vaccination

Vaccination was another key element in the UK pandemic response, and national governments consequently encouraged workplaces to support their staff in taking up COVID-19 vaccines (38). In this survey, providing time off work was the most common method of promoting vaccination across occupational groups (predicted probability range 50-86%) and use of promotional materials was also relatively common (22-78%), likely reflecting these nationally-recommended strategies. Providing vouchers or other incentives, which was at the employers discretion and not explicitly recommended in national guidance, was very uncommon across occupations (0-6%). Vaccination was never mandatory for the general public in England or Wales and mandatory vaccination policies were uncommon across most occupations except health and social care. Relatively high reporting in these groups may reflect mandates in England on two-dose vaccination for frontline care home staff (November 2021-March 2022) (39) and planned mandates for all frontline staff working in health and social care environments that were revoked in January 2022(39). High reported usage of other promotion strategies within this group likely reflect the impact of national prioritisation of these occupational groups for vaccination (40).

Occupations with the lowest level of workplace promotions – e.g., tradespeople, and transport and mobile machine operatives – were also those identified in previous studies as having lower vaccine uptake(24,41,42). A direct relationship between workplace promotion of vaccination and vaccine uptake cannot be inferred from this analysis, but is plausibly one of many work-related and wider sociodemographic factors influencing uptake in these groups. Investigation into effective workplace support and promotion of vaccination in these occupations may help to strengthen uptake in the event of future waves of COVID-19 or other vaccine-preventable outbreaks.

#### Workers’ Perceptions of Mitigation Methods

During both a period of stringent restrictions (third national lockdown) and a period of less intense restrictions (late February 2022), the majority of respondents across all occupational groups (>50%) agreed or strongly agreed that most work-related mitigations were reasonable and worthwhile. While proportional agreement was lower in February 2022, this trend remained in place – reflecting ongoing willingness to implement workplace measures to address transmission even with the change in national-level regulations. Patterns of agreement appeared relatively consistent across occupations, with agreement tending to be lower for physical distancing than other measures. While there was relatively high cross-occupational agreement for most measures, patterns of reporting that a given mitigation was not relevant or possible varied in line with likely job roles, and was prominent in trade and transport occupations.

While this sample of workers may be impacted by self-selection towards those with a high-level of concern about controlling the spread of COVID-19, it provides valuable multi-occupational insight into workers’ views about mitigations. The underlying attitudinal determinants and impact of these perceptions were beyond the scope of this survey. Workers may be more likely to adhere to public health interventions that they feel are reasonable and worthwhile(43). Further investigation and continued communication between workers and those developing workplace guidelines could strengthen understanding and implementation of mitigations for COVID-19 and in future public health emergencies.

### Strengths and Limitations

Strengths of this analysis included the large online cohort that enabled a multi-occupational investigation into the implementation and perception of work-related mitigation methods during the COVID-19 pandemic. This analysis also covered phases of the pandemic beyond the first and second wave, and these periods are currently underrepresented in the literature around occupation and COVID-19 and involve significant change in restrictions in England and Wales.

This analysis has a number of limitations as a result of its design and delivery. The subsample of respondents was not representative of the English and Welsh population, containing a high proportion of older and clinically vulnerable workers. Further, although the survey allowed investigation into mitigation methods over several key time periods, it was deployed at a single timepoint in February 2022. Consequently, responses may have been affected by recall bias – particularly for earlier timepoints. To reduce the risk of recall bias, relatively few key timepoints comprising key periods of restrictions were selected. Findings may also not be generalisable to earlier or later time periods outside of those investigated, though may be similar during periods of comparable restrictions.

To reduce burden, not all items were investigated over time and some pertained to whether a given strategy had ever been used during the whole survey period. This potentially masked important between-occupational differences in the persistence and implementation of given methods. Due to burden-related limitations, items tended to focus on presence of rather than adherence to work-related mitigations and were limited in detail. Many items were measured using broad ordinal scales to simplify survey delivery. Mitigation methods could not be directly linked to clinical outcomes due to data-related limitations, including a lack of adequate power and appropriate measures to investigate this relationship in the relevant timepoints.

Responses may have also been influenced by social desirability bias, particularly where behaviours were subject to national guidance or employer-level mandates. Participants’ behaviour in the workplace and perception of mitigations were also plausibly influenced by self-selection of cohort participants with a high degree of interest and motivation around preventing COVID-19; the impact on estimates of between-occupational differences is unknown. Responses may also have been affected by nation (England versus Wales), but the sample size did not allow stratification by region. Guidance was similar across both nations during most survey periods. In Wales but not England, guidance to wear face masks in some public spaces will still in place at the time of the survey and may have impacted related responses.

## Conclusions

Risk-relevant workplace features and mitigation methods differed substantially between occupations and over time during the third and fourth pandemic waves in the UK. Between-occupational differences corresponded to occupational variation in workplace environment, job roles, and legislation and guidance. Across occupations, there was a tendency towards reduced mitigations during periods of less intense national restrictions on social mixing. However, workers appeared to have a high level of agreement with most mitigation methods in the workplace, even after the relaxation of most national-level restrictions. Further investigation into effective workplace support for flagship national mitigation initiatives, such as regular antigen testing and promotion of vaccination, may be warranted to inform future pandemic planning.

## Supporting information

STROBE Checklist

## Data Availability

We aim to share aggregate data from this project on our website and via a "Findings so far" section on our website - https://ucl-virus-watch.net/. We also share some individual record level data on the Office of National Statistics Secure Research Service. In sharing the data we will work within the principles set out in the UKRI Guidance on best practice in the management of research data. Access to use of the data whilst research is being conducted will be managed by the Chief Investigators (ACH and RWA) in accordance with the principles set out in the UKRI guidance on best practice in the management of research data. We will put analysis code on publicly available repositories to enable their reuse.

## Interests

AH serves on the UK New and Emerging Respiratory Virus Threats Advisory Group and is a member of the COVID-19 transmission sub-group of the Scientific Advisory Group for Emergencies (SAGE).

## Footnotes

1 The COVID-19 Job Exposure Matrix is a six-dimension measure classifying occupational risk of SARS-CoV-2 transmission based on a range of workplace features. Please see (26) for more details.

