## Supplementary Materials for "Between-Occupation Differences in Work-Related COVID-19 Mitigation Strategies over Time: Analysis of the Virus Watch Cohort in England and Wales"

### Supplementary Table 1. UK Standard Occupational Classification 2020 (SOC-2020) Codes within Virus Watch Occupational Categories

| Virus Watch Occupational Group | Three Most Prevalent Occupations per Group* - UK Standard Occupation Classification 2020 Unit Groups |
|---|---|
| Administrative & Secretarial Occupations | 1. Other administrative occupations n.e.c. (25%, n= 216)<br>2. Book-keepers, payroll managers, and wage clerks (11%, n=94)<br>3. Office managers (7%, n=62) |
| Healthcare Occupations | 1. Other nursing professionals (26%, n=153)<br>2. Generalist medical practitioners (8.2%, n=48)<br>3. Community nurses (5.5%, n=32) |
| Indoor Trades, Process & Plant Occupations | 1. Warehouse operatives (8.5%, n=35)<br>2. Metalworking production and maintenance fitters (8.3%, n=34)<br>3. Electricians and electrical fitters (7.5%, n=31) |
| Leisure & Personal Service Occupations | 1. Cleaners and domestics (15%, n=46)<br>2. Kitchen and catering assistants (7.2%, n=22)<br>3. Waiters and waitresses (6.9%, n=21) |
| Managers, Directors & Senior Officials | 1. Financial managers and directors (18%, n=93)<br>2. Managers and directors in retail and wholesale (10%, n=52)<br>3. Property, housing, and estate managers (8.6%, n=44) |
| Other Professionals & Associate Professionals | 1. Programmers and software development professionals (5.7%, n=111)<br>2. Management consultants and business analysts (4.1%, n=79)<br>3. Chartered and certified accountants (3.9%, n=75) |
| Outdoor Trade Occupations | 1. Gardeners and landscape gardeners (23%, n=44)<br>2. Construction and building trades n.e.c. (15%, n=28) |

| | |
|---|---|
| | 3. Farmers (12%, n=23) |
| Sales & Customer Service Occupations | 1. Sales and retail assistants (40%, n=117)<br>2. Customer service occupations n.e.c. (12%, n=36)<br>3. Retail cashiers and check-out operators (11%, n=31) |
| Social Care & Community Protective Services | 1. Care workers and home carers (27%, n=98)<br>2. Welfare and housing associate professionals n.e.c. (12%, n=46)<br>3. Clergy (8.2%, n=30) |
| Teaching, Education & Childcare Occupations | 1. Higher education teaching professionals (16%, n=103)<br>2. Secondary education teaching professionals (12%, n=80)<br>3. Teaching assistant (12%, n=78) |
| Transport & Mobile Machine Operatives | 1. Large goods vehicle drivers (19%, n=26)<br>2. Road transport drivers n.e.c. (17%, n=23)<br>3. Delivery drivers and couriers (16%, n=22) |

**Abbreviations:** n.e.c. = not elsewhere classified; * Limited to three most prevalent occupations per category to prevent declarative disclosure and due to large number of occupations across sample (*n* =412). Occupations defined based on four-digit UK Standard Occupational Classification codes

### Virus Watch Work-Related Mitigations Survey (February 2022)

**Your Experiences at Work During the Pandemic**

If you have been employed or self-employed during the pandemic, we would like to ask some questions about your work-related experiences.

We understand that people's working situations can vary considerably and can be difficult to capture in a questionnaire. There are no right or wrong answers so please just let us know about your situation.

1) **Which of the following descriptions best applies to your employment situation during the following periods?** If your employment status changed during any period, please select the description with the longest hours

| | |
|---|---|
| Late December 2020 – March 2021 (third lockdown) | Not working (e.g. unemployed, retired) / On full-time furlough / Working up to 20 hours per week / Working 20-35 hours per week / Working more than 35 hours per week |
| July – December 2021 (restrictions lifted) | Not working (e.g. unemployed, retired) / On full-time furlough / Working up to 20 hours per week / Working 20-35 hours per week / Working more than 35 hours per week |
| Late December 2021 – January 2022 (Omicron/Phase 2 restrictions) | Not working (e.g. unemployed, retired) / On full-time furlough / Working up to 20 hours per week / Working 20-35 hours per week / Working more than 35 hours per week |
| Current | Not working (e.g. unemployed, retired) / On full-time furlough / Working up to 20 hours per week / Working 20-35 hours per week / Working more than 35 hours per week |

2) *[If working full time/ part time at any point]* **Were you working in the same job or similar jobs/roles throughout?** Yes/ No

3) [If yes]: **What is/was your job? If you work(ed) in multiple jobs at the same time, please provide answers for the position that you would consider your 'main' job.** [Text]

4) *[If working full time/ part time at any point]*: **During the following period(s), how many days did you work <u>outside your home</u> in an average week?** Please round down to the nearest day (e.g. if you worked two and a half days, choose two)

| | |
|---|---|
| Late December 2020 – March 2021 (third lockdown) | None / Half a day / 1 day / 2 days / 3 days / 4 days/ 5 or more days |
| July – December 2021 (restrictions lifted) | None / Half a day / 1 day / 2 days / 3 days / 4 days/ 5 or more days |
| Late December 2021 – January 2022 (Omicron/Phase 2 restrictions) | None / Half a day / 1 day / 2 days / 3 days / 4 days/ 5 or more days |
| Current | None / Half a day / 1 day / 2 days / 3 days / 4 days/ 5 or more days |

5) *[If attended workplace at any point]* **Please select the description of your main working environment that best applies:** Working (mostly) outside/ Working partly inside / Working (mostly) inside

6) *[If works indoors at least sometimes]:* **Does your workplace regularly use any of the following ventilation methods? Please select all that apply:**

| | |
|---|---|
| Opening doors and/or windows when weather allows | Yes/ No / Don't know/ NA |
| Mechanical ventilation system | Yes/ No / Don't know/ NA |
| Air purifiers/cleaners (e.g. HEPA filter) | Yes/ No / Don't know/ NA |

7) *[For periods when attended workplace]* **During a typical workday, how many people did you share your workspace with across the whole day (even if socially distanced)?** Please provide your best estimate of the number of people you shared the workspace with across a typical day, including co-workers and customers/client/patients (if applicable).

| | |
|---|---|
| Late December 2020 – March 2021 (third lockdown) | None / Fewer than 10 people / 10-30 people/ More than 30 people |
| July – December 2021 (restrictions lifted) | None / Fewer than 10 people / 10-30 people/ More than 30 people |
| Late December 2021 – January 2022 (Omicron/Phase 2 restrictions) | None / Fewer than 10 people / 10-30 people/ More than 30 people |
| Current | None / Fewer than 10 people / 10-30 people/ More than 30 people |

8) *[If attended workplace at any point]* **Please indicate whether your workplace ever used any of the following measures to improve people's ability to social distance / to reduce contact:**

- Limiting the number of people on the premises: Yes / No / Don't know / NA
- Staggering the timing of breaks: Yes / No / Don't know / NA
- Staggering shifts: Yes / No / Don't know / NA
- Workplace bubbles (i.e. only working with a small, consistent group of colleagues): Yes / No / Don't know / NA
- One-way systems on the premises: Yes / No / Don't know / NA
- Reconfiguring the space (e.g. putting more space between tables or desks): : Yes / No / Don't know / NA
- Using posters, floor markings, or reminders to keep social distance: Yes / No / Don't know / NA
- Using screens or barriers: Yes / No / Don't know / NA

9) *[For periods when workspace was shared]* **Typically, to what degree could social distance (2m+) be maintained from other people in the same work area (e.g., colleagues, the public, patients)**

| | |
|---|---|
| Late December 2020 – March 2021 (third lockdown) | Never/ Rarely/ Sometimes / Most of the Time/ Always |
| July – December 2021 (restrictions lifted) | Never/ Rarely/ Sometimes / Most of the Time/ Always |
| Late December 2021 – January 2022 (Omicron/Phase 2 restrictions) | Never/ Rarely/ Sometimes / Most of the Time/ Always |
| Current | Never/ Rarely/ Sometimes / Most of the Time/ Always |

10) *[If attended workplace at any point]* **Typically at work, how often do you touch surfaces or other things that multiple people may also touch or use (e.g. tabletops, handrails, cash)?** Never/ Very infrequently/ Frequently/ Very frequently/Don't know

11) *[If surfaces ever shared]* **In general, how frequently were surfaces (e.g. tabletops, handrails) that are regularly touched by other people cleaned at your workplace?**

| | |
|---|---|
| Late December 2020 – March 2021 (third lockdown) | Never/ Very infrequently/ Frequently/ Very frequently/Don't know |
| July – December 2021 (restrictions lifted) | Never/ Very infrequently/ Frequently/ Very frequently/Don't know |
| Late December 2021 – January 2022 (Omicron/Phase 2 restrictions) | Never/ Very infrequently/ Frequently/ Very frequently/Don't know |
| Current | Never/ Very infrequently/ Frequently/ Very frequently/Don't know |

12) *[For periods when attended workplace]* **Typically, how frequently do you wash or sanitise your hands at work?**

| | |
|---|---|
| Late December 2020 – March 2021 (third lockdown) | Never / 1-5 times a day / 6-10 times a day / more than 10 times a day |
| July – December 2021 (restrictions lifted) | Never / 1-5 times a day / 6-10 times a day / more than 10 times a day |
| Late December 2021 – January 2022 (Omicron/Phase 2 restrictions) | Never / 1-5 times a day / 6-10 times a day / more than 10 times a day |
| Current | Never / 1-5 times a day / 6-10 times a day / more than 10 times a day |

13) *[For periods when workspace was shared]* **During a typical workday, how often were face coverings worn during the following periods?**

| | |
|---|---|
| Late December 2020 – March 2021 (third lockdown) | **By you:** Never/ Rarely/ Sometimes / Most of the Time/ Always<br><br>**By others:** Never/ Rarely/ Sometimes / Most of the Time/ Always |
| July – December 2021 (restrictions lifted) | **By you:** Never/ Rarely/ Sometimes / Most of the Time/ Always<br><br>**By others:** Never/ Rarely/ Sometimes / Most of the Time/ Always |
| Late December 2021 – January 2022 (Omicron/Phase 2 restrictions) | **By you:** Never/ Rarely/ Sometimes / Most of the Time/ Always<br><br>**By others:** Never/ Rarely/ Sometimes / Most of the Time/ Always |
| Current | **By you:** Never/ Rarely/ Sometimes / Most of the Time/ Always<br><br>**By others:** Never/ Rarely/ Sometimes / Most of the Time/ Always |

14) *[If workspace was shared]* **Did your workplace ever provide face coverings to reduce risk of transmission?**

| | |
|---|---|
| For workers | No / Yes/ Don't know / NA |
| For customers/clients/patients | No / Yes/ Don't know / NA |

15) *[If workspace was shared]*.**Typically, which of the following best describes how you spend breaktimes / lunchtime?** Alone / With others outdoors / With others indoors

16) *[For periods when workspace was shared]* **During the following periods, do you think that people in your workplace tended to be less likely to practice protective measures (e.g. social distancing, wearing face coverings) during rest periods or breaks?**

| | |
|---|---|
| Late December 2020 – March 2021 (third lockdown) | Yes/ No/ Don't know/ NA |
| July – December 2021 (restrictions lifted) | Yes/ No/ Don't know/ NA |
| Late December 2021 – January 2022 (Omicron/Phase 2 restrictions) | Yes/ No/ Don't know/ NA |
| Current | Yes/ No/ Don't know/ NA |

17) *[For periods when workspace was shared]* **Did your workplace organise social events outside of working hours or allow social events on work premises including food and/or drinks (i.e. parties)?**

| | |
|---|---|
| Late December 2020 – March 2021 (third lockdown) | Never / Rarely / Sometimes / Frequently / NA |
| July – December 2021 (restrictions lifted) | Never / Rarely / Sometimes / Frequently / NA |
| Late December 2021 – January 2022 (Omicron/Phase 2 restrictions) | Never / Rarely / Sometimes / Frequently / NA |
| Current | Never / Rarely / Sometimes / Frequently / NA |

18) *[For periods when attended workplace]* **Did your workplace require or recommend regular (e.g. twice weekly) COVID-19 lateral flow tests (swab tests where you get the result with 30 minutes) to attend work at any time during the following periods?**

| | |
|---|---|
| Late December 2020 – March 2021 (third lockdown) | Required/Recommended/Not discussed/Don't know |
| July – December 2021 (restrictions lifted) | Required/Recommended/Not discussed/Don't know |
| Late December 2021 – January 2022 (Omicron/Phase 2 restrictions) | Required/Recommended/Not discussed/Don't know |
| Current | Required/Recommended/Not discussed/Don't know |

19) *[If attended workplace at any point]* **Did your workplace ever provide lateral flow testing?**

| | |
|---|---|
| On-site testing | Yes/ No/ Don't know |
| At-home test kits | Yes/ No/ Don't know |

20) **How did your workplace promote COVID-19 vaccination, if at all. Please select all that apply:**

| | |
|---|---|
| Allowed time off work to get vaccinated | Yes/ No / Don't know |
| Promotional materials (e.g. posters, staff emails) | Yes/ No / Don't know |
| Mandatory vaccination for work | Yes/ No / Don't know |
| Vouchers (or similar) for getting vaccinated | Yes/ No / Don't know |
| Other | Yes/ No / Don't know |

21) **At <u>this time last year</u> (i.e., third national lockdown, prior to widespread vaccination of adults), how much do you agree that the following measures would have been reasonable and worthwhile to prevent COVID-19 transmission at your work?** We are interested in your opinions, even if the measures were not actually implemented at your work

| | Strongly agree (very reasonable and worthwhile) | Agree | Neither agree nor disagree | Disagree | Strongly disagree (not at all reasonable or worthwhile) | Not possible/relevant in my job |
|---|---|---|---|---|---|---|
| Working from home when possible | | | | | | |
| Regular Covid-19 tests for workers | | | | | | |
| Proof of vaccination required for workers | | | | | | |
| Proof of vaccination required for customers/clients | | | | | | |
| Physical distancing (e.g., 2m or limiting number of customers) | | | | | | |
| Workplace well-ventilated (e.g., opening doors/windows, ventilation system / air purification system) | | | | | | |
| Face coverings required for workers | | | | | | |
| Face coverings required for customers/clients/patients | | | | | | |
| Using screens or barriers | | | | | | |
| Regular cleaning of surfaces | | | | | | |

22) **At <u>current stage</u> of the pandemic, how much do you agree that the following measures would be reasonable and worthwhile to prevent COVID-19 transmission at your work?** We are interested in your opinions, even if the measures were not actually implemented at your work

| | Strongly agree (very reasonable and worthwhile) | Agree | Neither agree nor disagree | Disagree | Strongly disagree (not at all reasonable or worthwhile) | Not possible/relevant in my job |
|---|---|---|---|---|---|---|
| Working from home when possible | | | | | | |
| Regular Covid-19 tests for workers | | | | | | |
| Proof of vaccination required for workers | | | | | | |
| Proof of vaccination required for customers/clients | | | | | | |
| Physical distancing (e.g., 2m or limiting number of customers) | | | | | | |
| Workplace well-ventilated (e.g., opening doors/windows, ventilation | | | | | | |

| |
|---|
| system/ air purification system) |
| Face coverings required for workers |
| Face coverings required for customers/clients/patients |
| Using screens or barriers |
| Regular cleaning of surfaces |

### Supplementary Table 2. Focus and Source for Items in Work-Related Mitigations Survey

| Item | Topic | Aim | Source |
|---|---|---|---|
| 1 | Employment status over time | Establishing eligibility | Virus Watch registration survey |
| 2 | Same job over time | Establishing eligibility | Created for survey |
| 3 | Job title | Establishing occupation (exposure) | Virus Watch registration survey |
| 4 | Days worked outside home | Screening for subsequent workplace related questions / potential exposure | Created for survey |
| 5 | Main working environment | Assess potential exposure (for ventilation item) | Adapted from COVID-19 JEM (1) |
| 6 | Ventilation methods | Assess mitigation - ventilation (aerosol transmission) | Created for survey |
| 7 | People in shared workspace | Assess potential exposure (for social distancing and other items) | Adapted from COVID-19 JEM |
| 8 | Methods to promote social distancing/reduce contact | Assess mitigation - social distancing (aerosol and droplet transmission) | Created for survey |
| 9 | Degree that social distance could be maintained | Assess mitigation - social distancing (aerosol and droplet transmission) | Adapted from COVID-19 JEM |
| 10 | Touching shared surfaces/objects | Assess potential exposure (for hygiene items) | Adapted from COVID-19 JEM |
| 11 | Frequency of surface/object cleaning | Assess mitigation – surface hygiene (indirect contact transmission) | Created for survey |
| 12 | Frequency of hand hygiene at work | Assess mitigation – hand hygiene (indirect/direct contact transmission) | Flu Watch Study (2) |
| 13 | Frequency of wearing face coverings | Assess mitigation – face coverings (primarily droplet transmission) | Adapted from COVID-19 JEM and Virus Watch contact survey (3) |
| 14 | Workplace provision of face coverings | Assess mitigation – face coverings (primarily droplet transmission) | Created for survey |
| 15 | How breaks/lunchtime spent | Assess potential exposure (for measures during breaks item) | Created for survey |
| 16 | Reduction in protective measures during breaks | Assess mitigation/exposure – stringency of measures during breaks (potentially all transmission pathways) | Created for survey |

| | | | |
|---|---|---|---|
| 17 | Work-related social events | Assess potential exposure – work-related social contact | Created for survey |
| 18 | Workplace LFT policy | Assess mitigation – case identification and isolation (all transmission pathways) | Created for survey |
| 19 | Workplace LFT provision | Assess mitigation – case identification and isolation (all transmission pathways) | Created for survey |
| 20 | Workplace COVID-19 vaccine promotion methods | Assess mitigation – reducing susceptibility esp. to severe outcomes | Created for survey |
| 21 | Perception of mitigation methods (third national lockdown) | Assess worker perceptions – period of high intensity of national measures | Created for survey |
| 22 | Perception of mitigation methods (February 2022) | Assess worker perceptions – period of low intensity of national measures | Created for survey |

**Abbreviations:** COVID-19 JEM – COVID-19 Job Exposure Matrix

**Supplementary Figure 1. Schematic Diagram of Virus Watch Mitigations Survey: Items included in analyses**

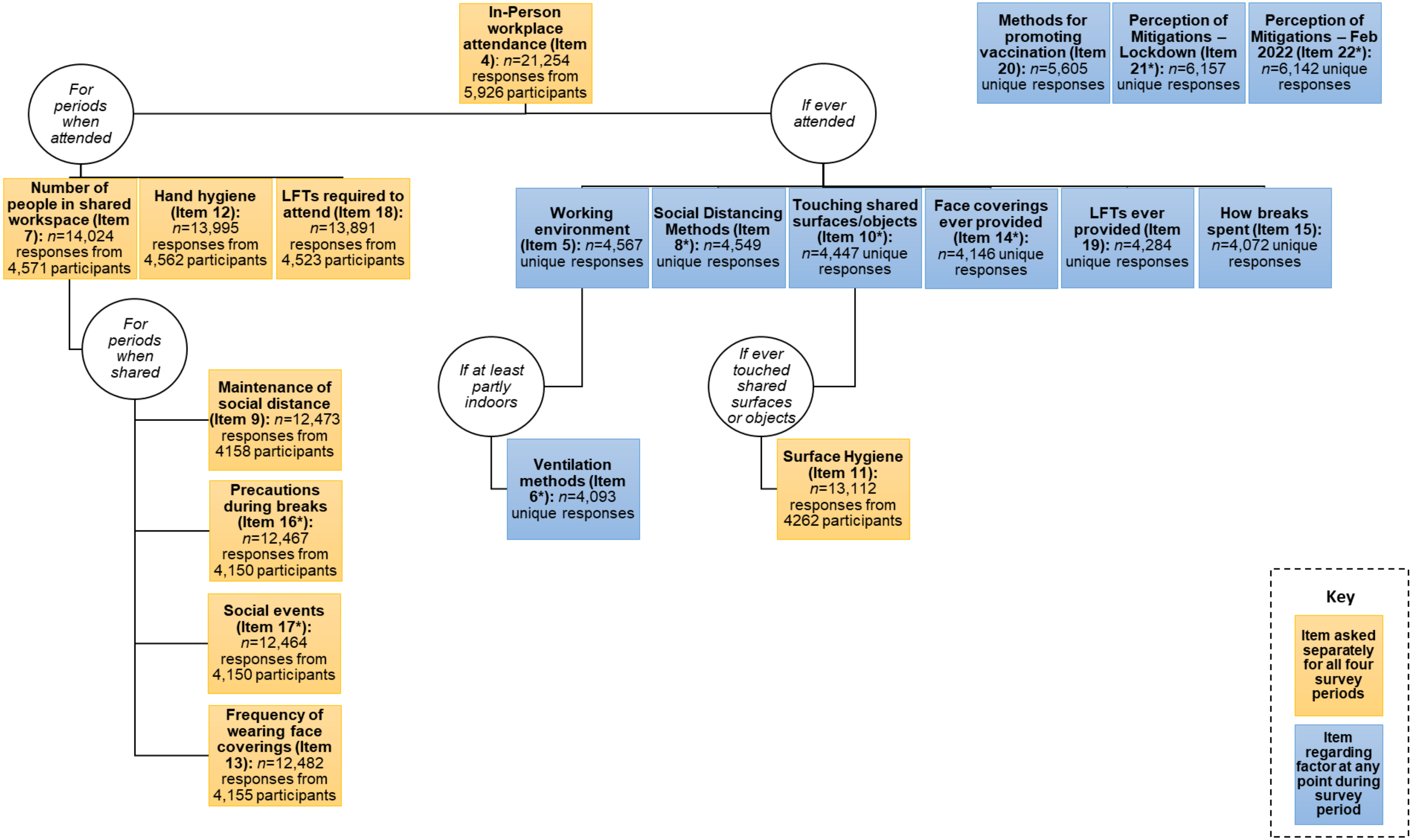

**Note:** Where item contained multiple sub-ratings, the overall maximum for the item is presented; ***** item contained 'Unsure/NA' responses, reported below

### Supplementary Table 3. 'Unsure'/'NA' Responses Excluded from Analyses for Relevant Items

| Item | Item Timeframe | Unsure/NA Responses |
|---|---|---|
| 6 | Whole survey period | *n*=381 for physical ventilation<br>*n*=678 for mechanical ventilation<br>*n*=1153 for filter ventilation |
| 8 | Whole survey period | *n*= 571 for limiting occupations<br>*n*= 743 for reconfiguring workspace<br>*n*= 1374 for staggering shifts<br>*n*= 740 for one-way systems<br>*n*= 735 for screens/barriers<br>*n*= 918 for workplace bubbles<br>*n*= 618 for posters/reminders<br>*n*= 1229 for staggering breaks |
| 10 | Whole survey period | *n*=40 |
| 14 | Whole survey period | *n*=316 for worker self-report<br>*n*=798 for other people on the worksite |
| 16 | Separate responses for all periods when workspace was shared | *n*=2571 responses for 951 participants |
| 17 | Separate responses for all periods when workspace was shared | *n*=1189 responses for 447 participants |
| 21 | Single timepoint (third national lockdown) | NA responses included in analyses due to different analytical method and aims – reported in main findings |
| 22 | Single timepoint (late February 2022) | NA responses included in analyses due to different analytical method and aims – reported in main findings |

**Note:** Items not included in this table did not have 'Unsure' or 'NA' response options; NA=not applicable

**Supplementary Figure 2. Flow Diagram of Participant Selection**

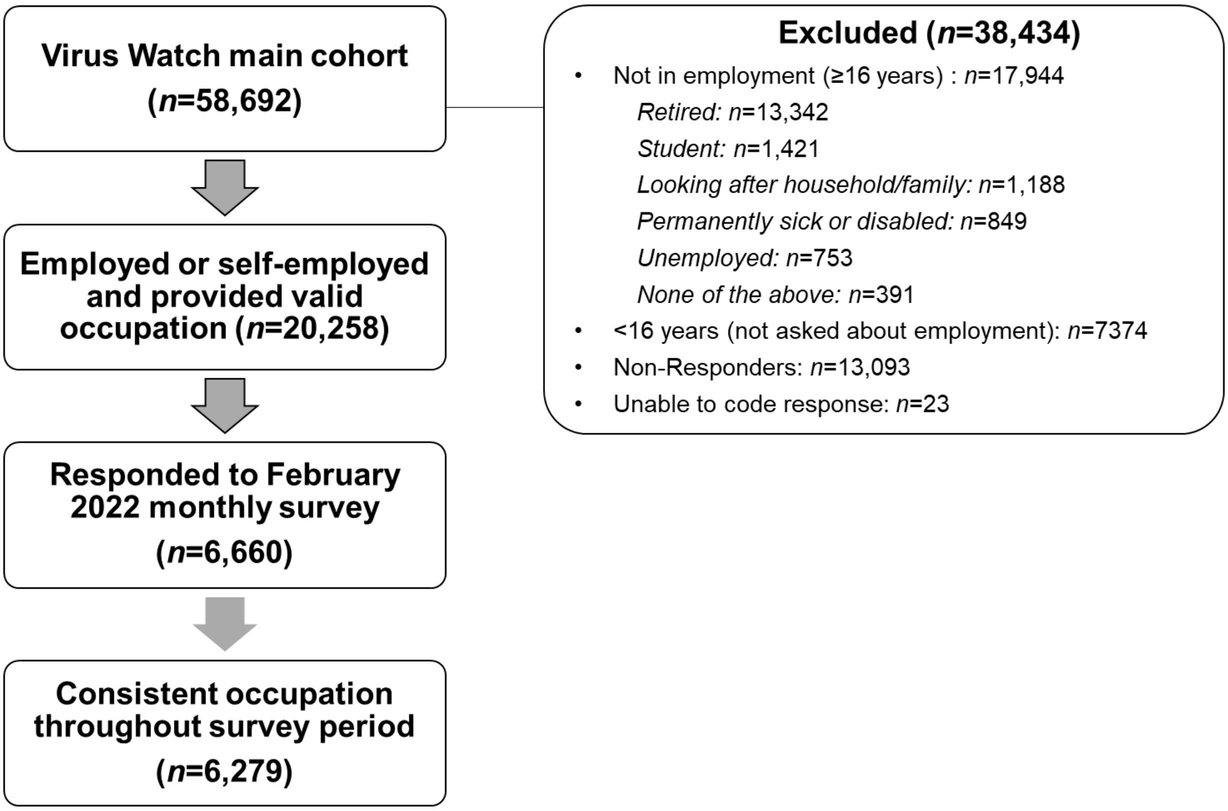

### Supplementary Table 4. Employment Status Across Survey Periods

| | **Late December 2020 – March 2021**, N = 6,225[1] | **July – December 2021**, N = 6,217[1] | **Late December 2021 – January 2022**, N = 6,206[1] | **Late February 2022**, N = 6,072[1] |
|---|---|---|---|---|
| Not working (e.g. unemployed, retired) | 476 (7.6%) | 337 (5.4%) | 479 (7.7%) | 496 (8.2%) |
| On full-time furlough | 347 (5.6%) | 70 (1.1%) | 40 (0.6%) | 18 (0.3%) |
| Working up to 20 hours per week | 1,273 (20%) | 1,468 (24%) | 1,437 (23%) | 1,371 (23%) |
| Working 20-35 hours per week | 1,217 (20%) | 1,353 (22%) | 1,317 (21%) | 1,312 (22%) |
| Working more than 35 hours per week | 2,912 (47%) | 2,989 (48%) | 2,933 (47%) | 2,875 (47%) |

[1]n (%)

**Supplementary Figure 3. Average Number of Days per Week Worked Outside the Home: Predicted probabilities by occupation over time (Adjusted Estimates)**

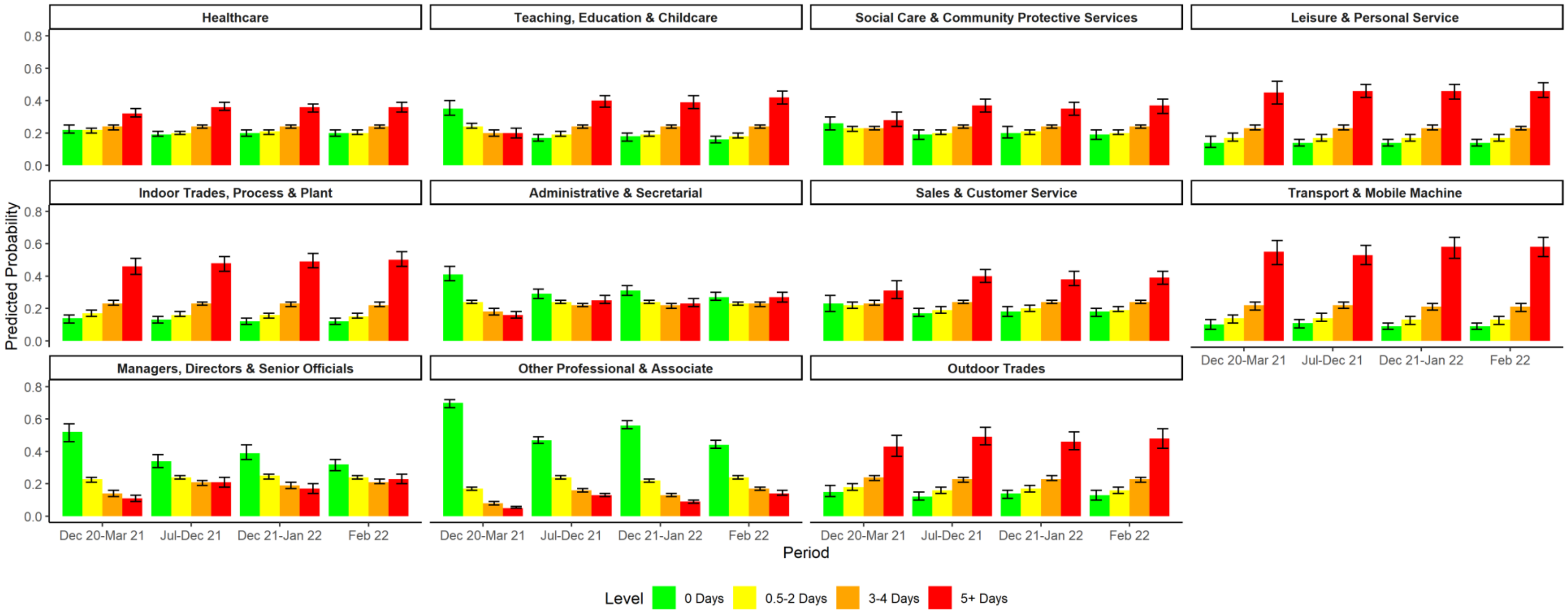

**Supplementary Figure 4. Average Number of Days per Week Worked Outside the Home, by Occupation and Time Period (Unadjusted Estimates)**

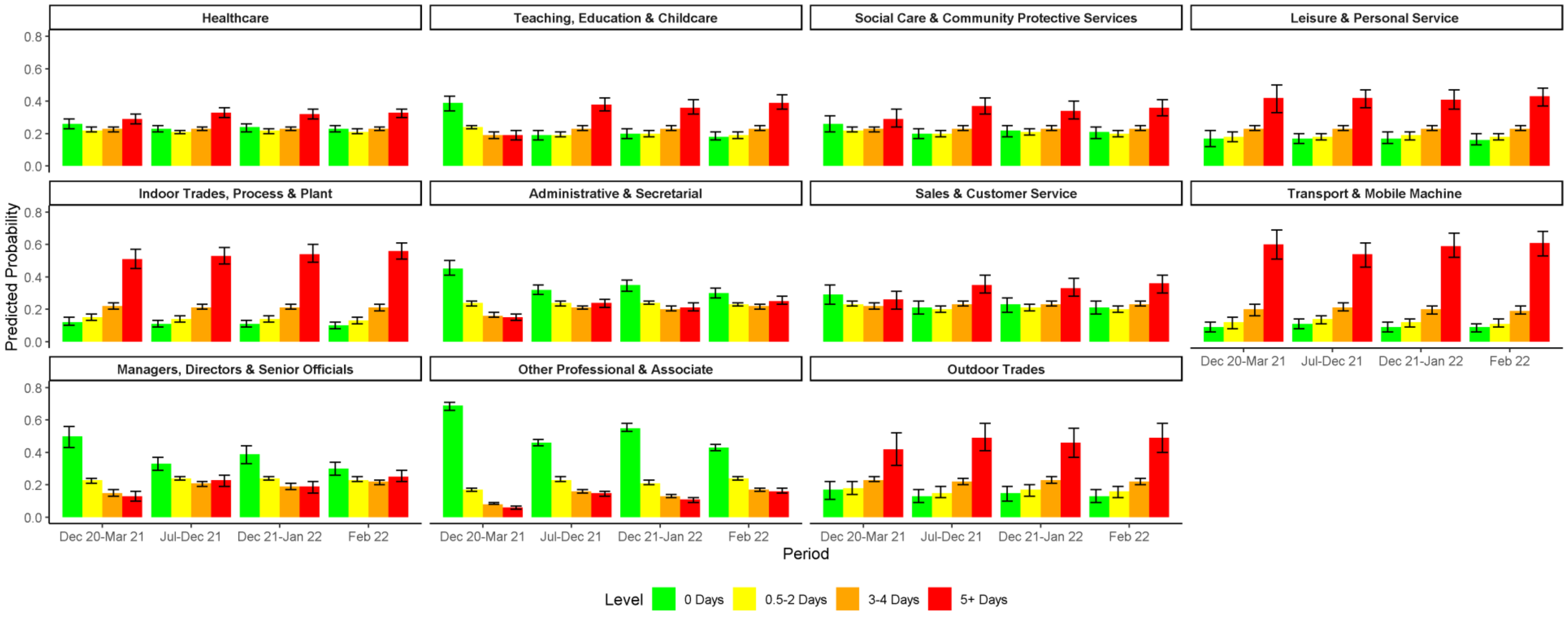

### Supplementary Figure 5. Number of People in Workspace across Day: Predicted Probabilities by occupation over time

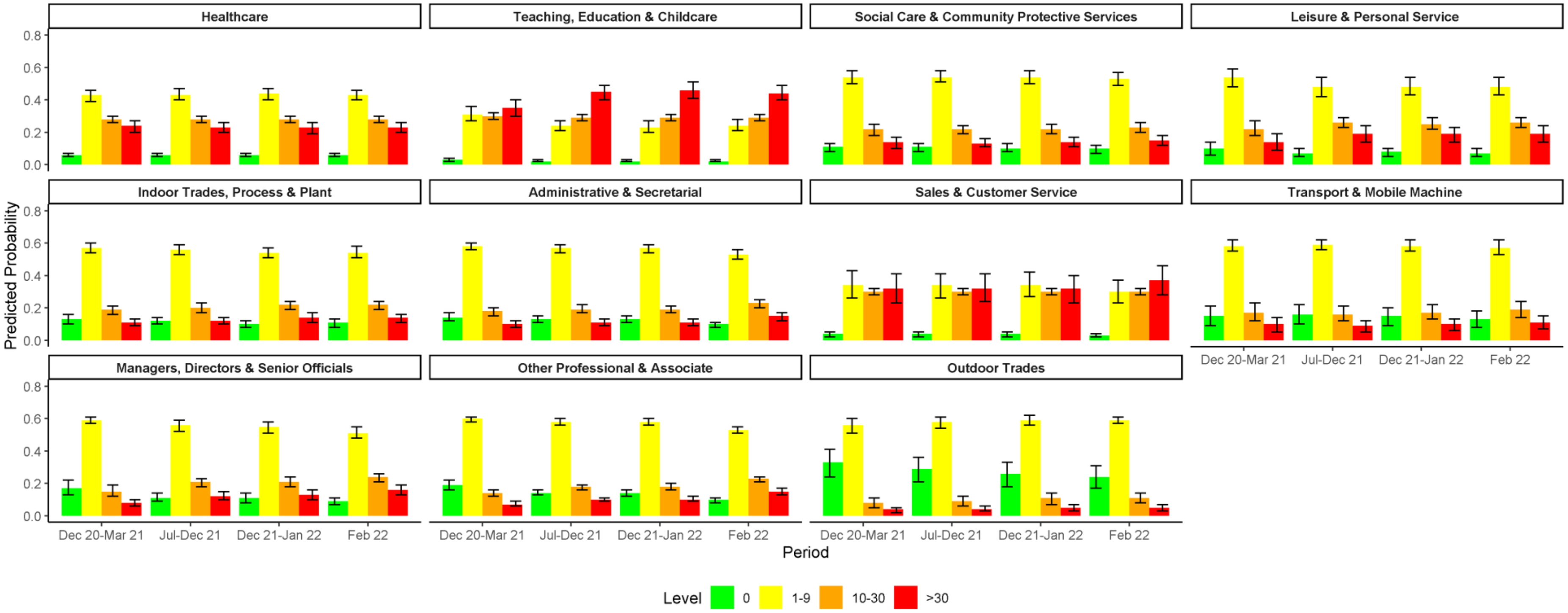

**Supplementary Figure 6. Extent to Which Social Distance Could be Maintained at Work: Predicted Probabilities by occupation over time**

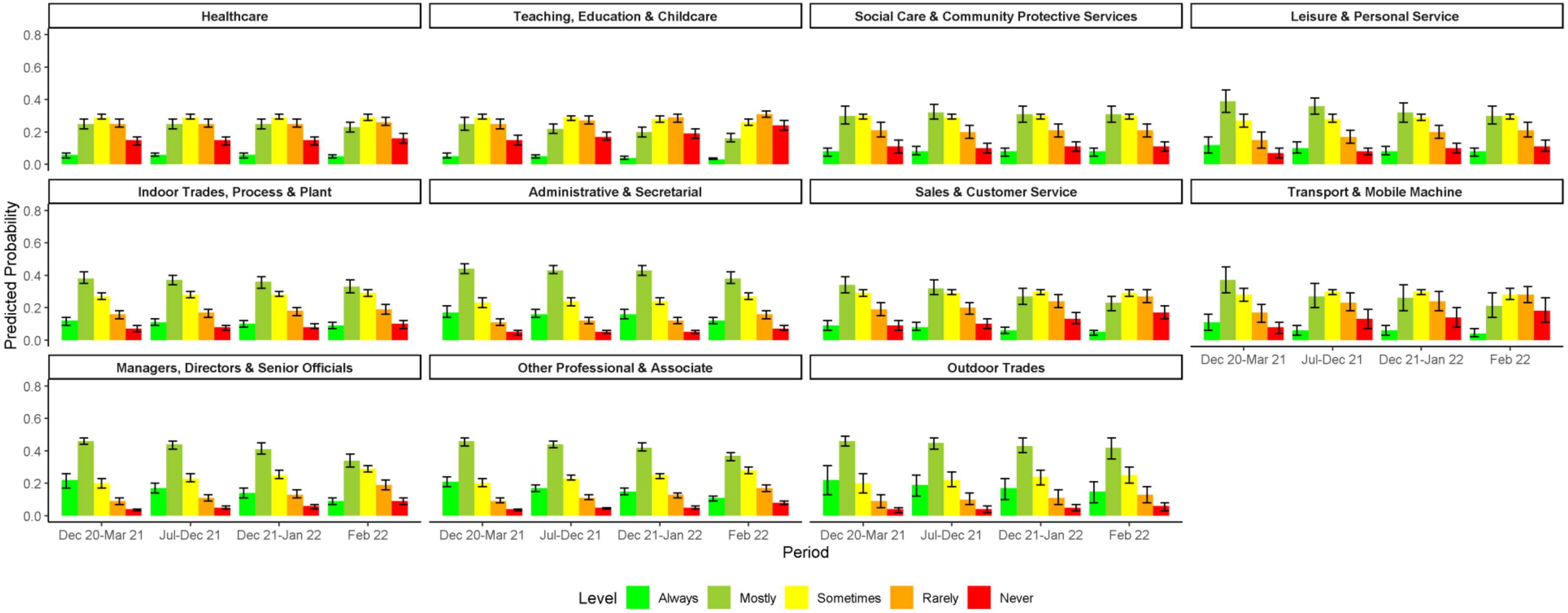

**Supplementary Figure 7. Working Environment: Predicted probabilities by occupation**

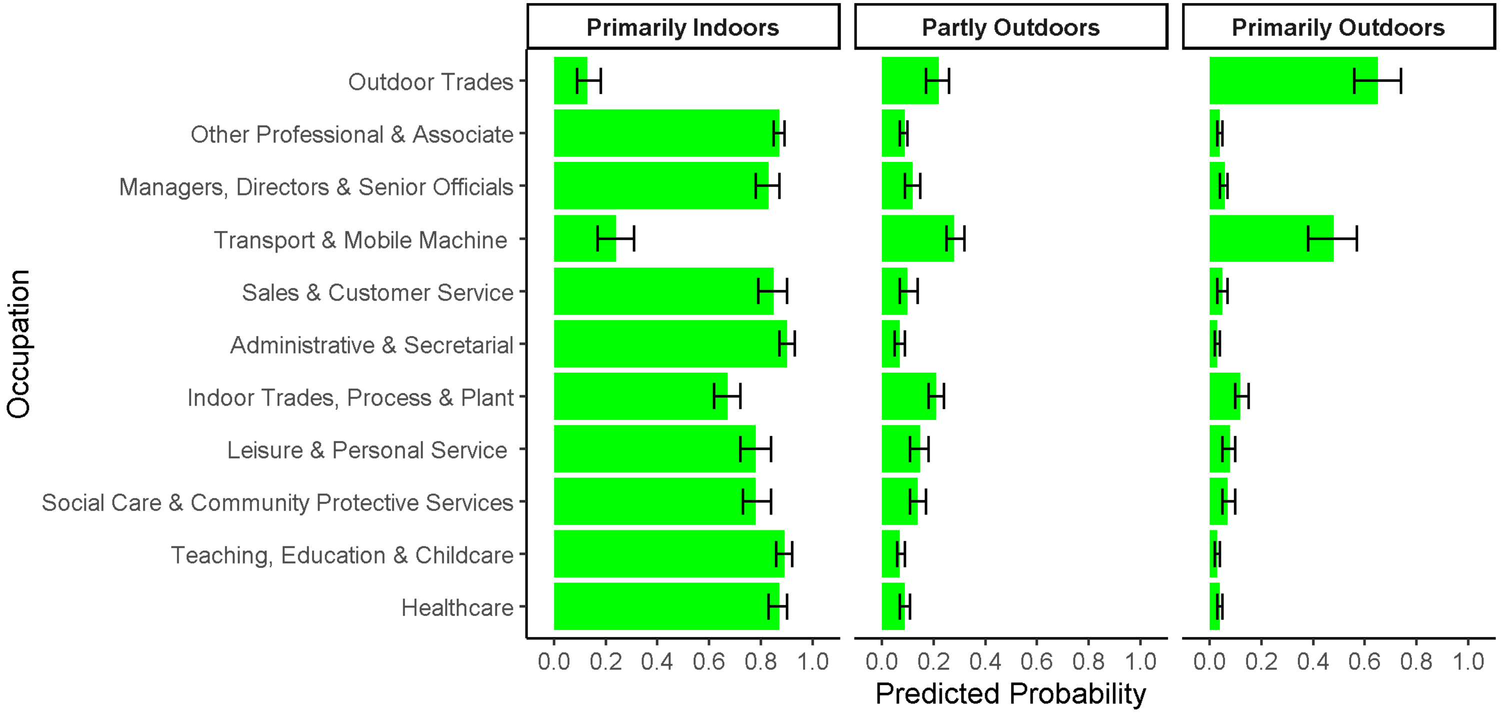

**Supplementary Figure 7. Ventilation Methods Used in the Workplace: Predicted probabilities by occupation**

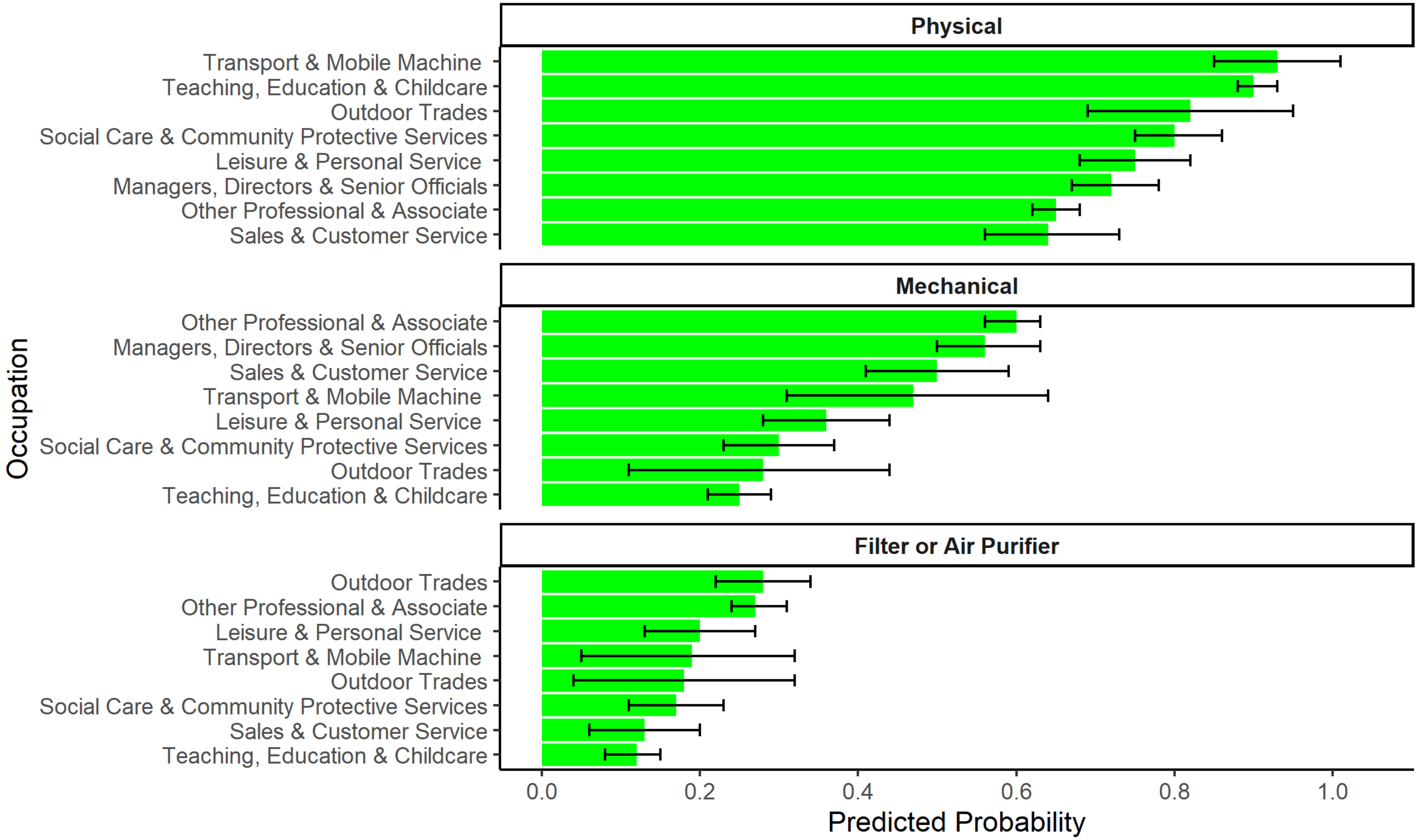

**Supplementary Figure 8. Frequency of Touching Shared Surfaces or Objects at Work: Predicted probabilities by occupation**

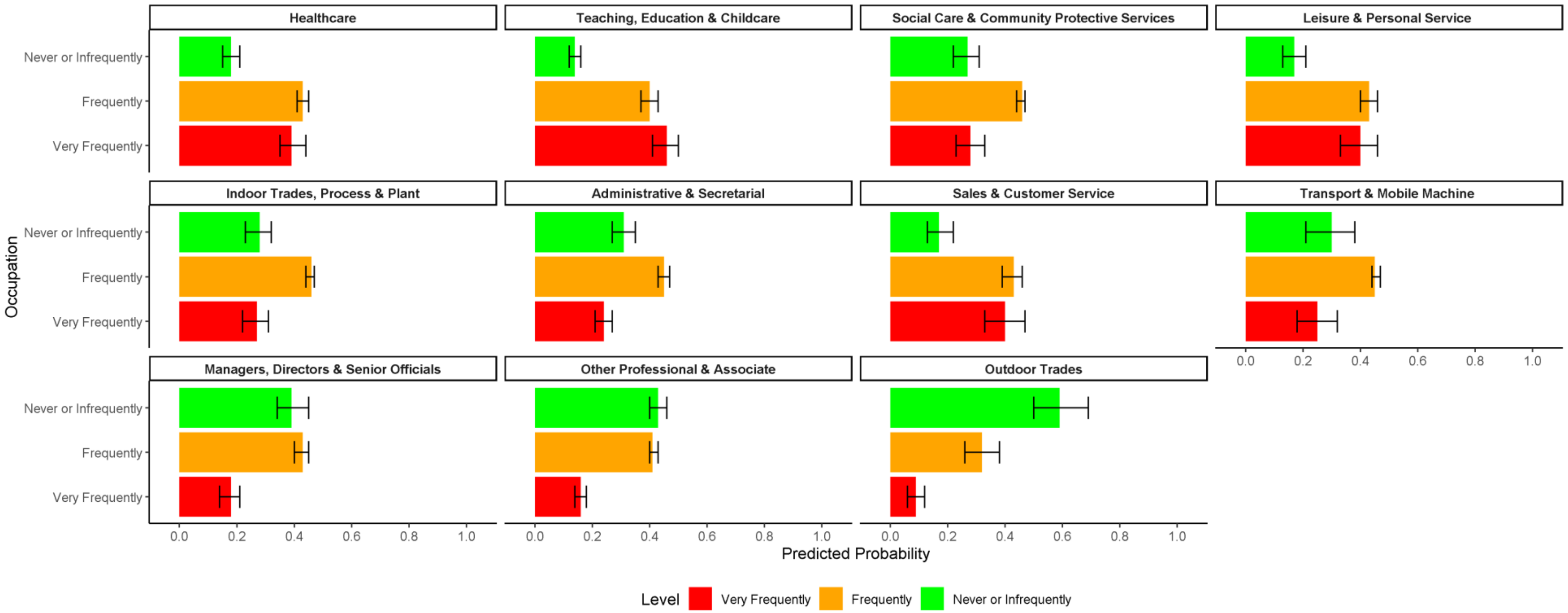

**Supplementary Figure 9.** Frequency of Surface Hygiene at Work: Predicted probabilities by occupation (a) and over time (b)

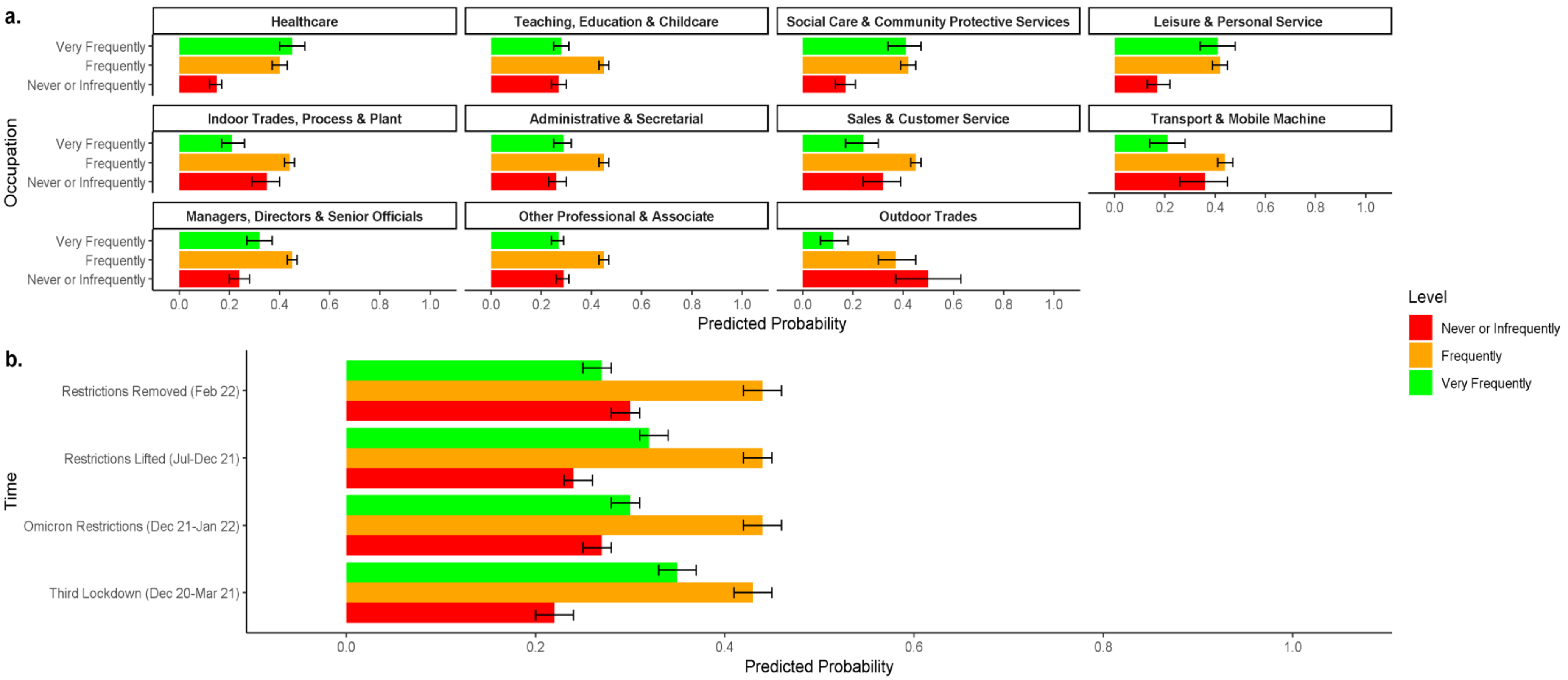

**Supplementary Figure 10. Workers' Personal Usage of Face Coverings: Predicted probabilities by occupation over time**

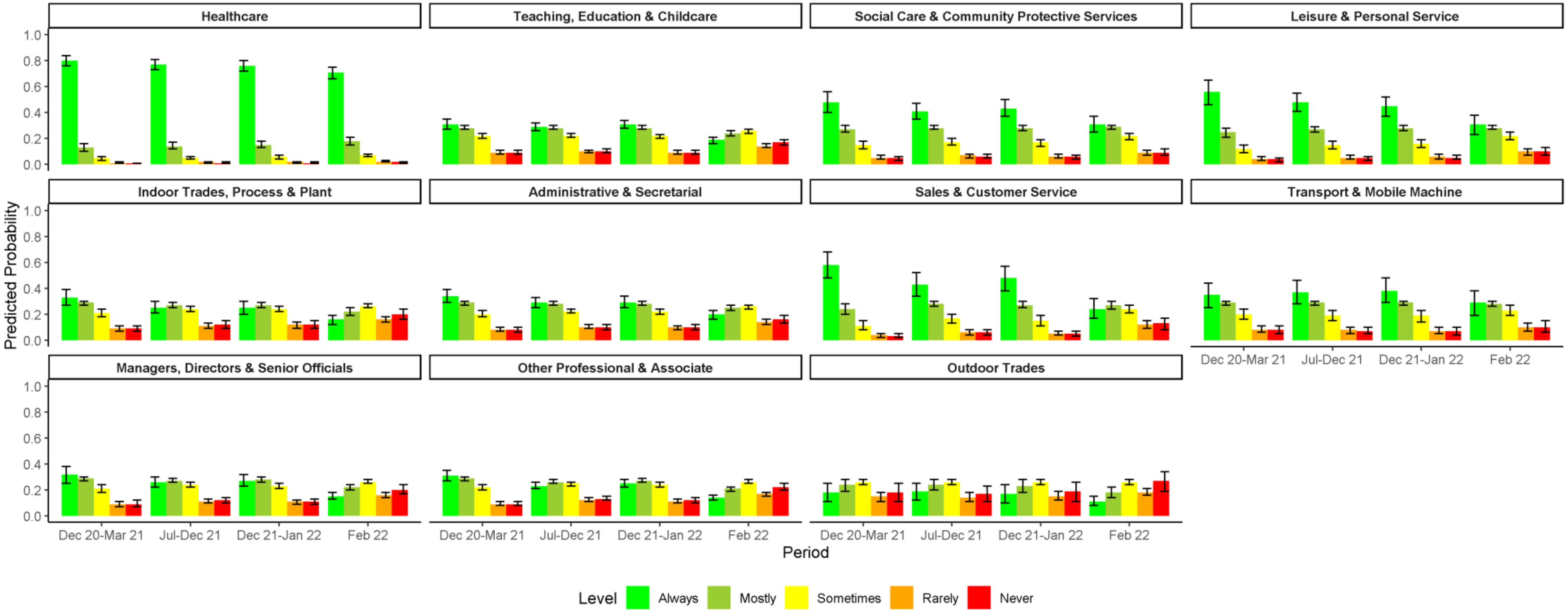

**Supplementary Figure 11. Usage of Face Coverings by Other People on the Worksite: Predicted probabilities by occupation over time**

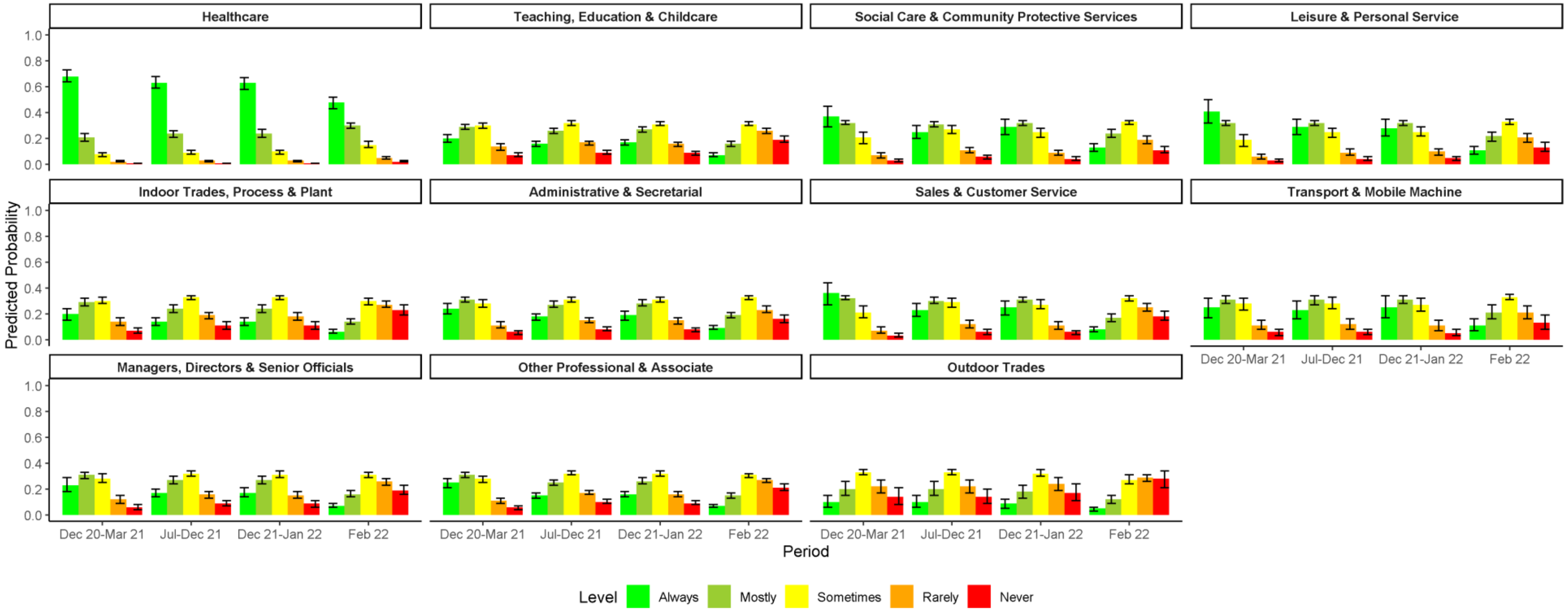

**Supplementary Figure 12. Workplace Provision of Face Coverings by Occupation**

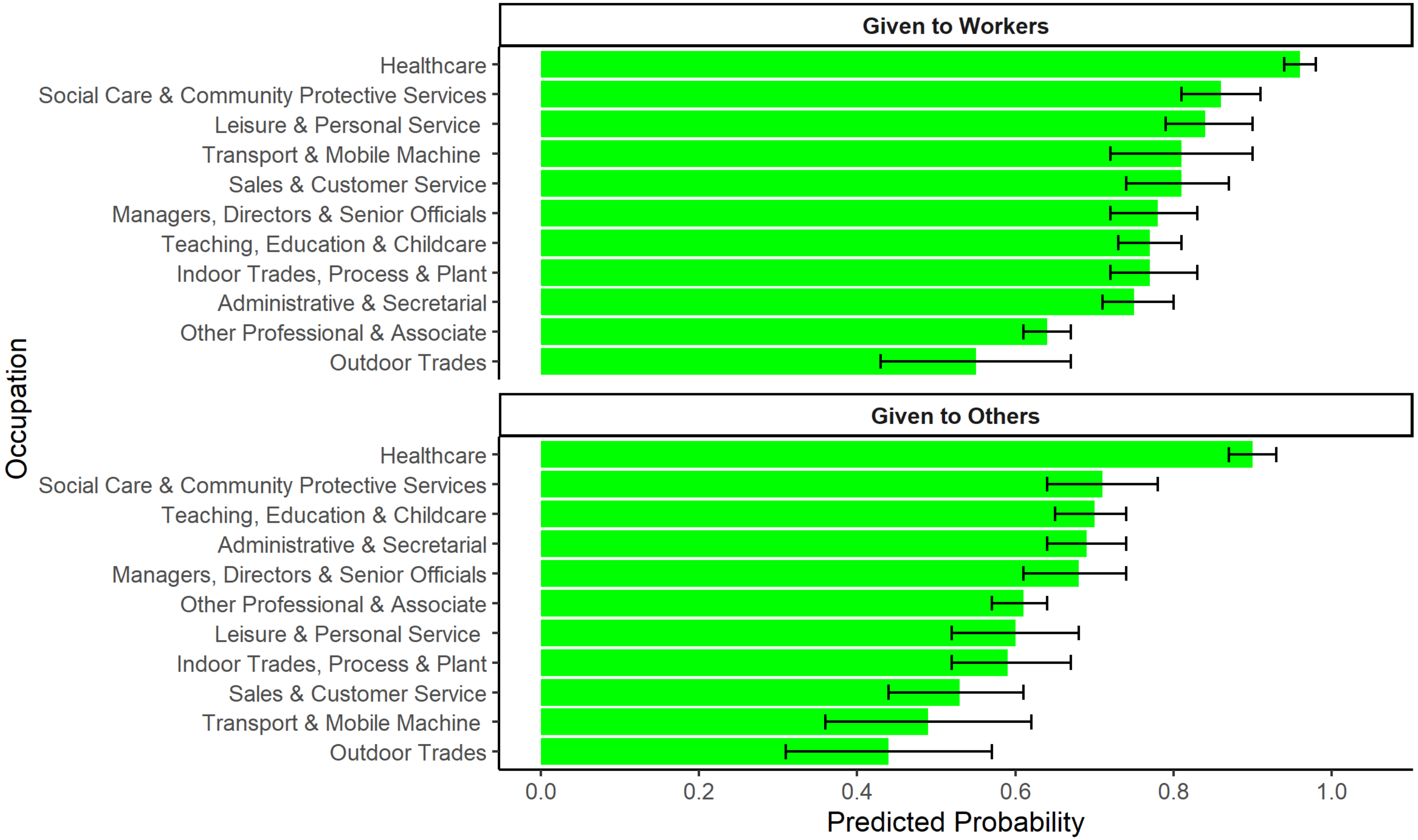

### Supplementary Figure 13. Typical Contact with Others during Breaks: Predicted probabilities by occupation

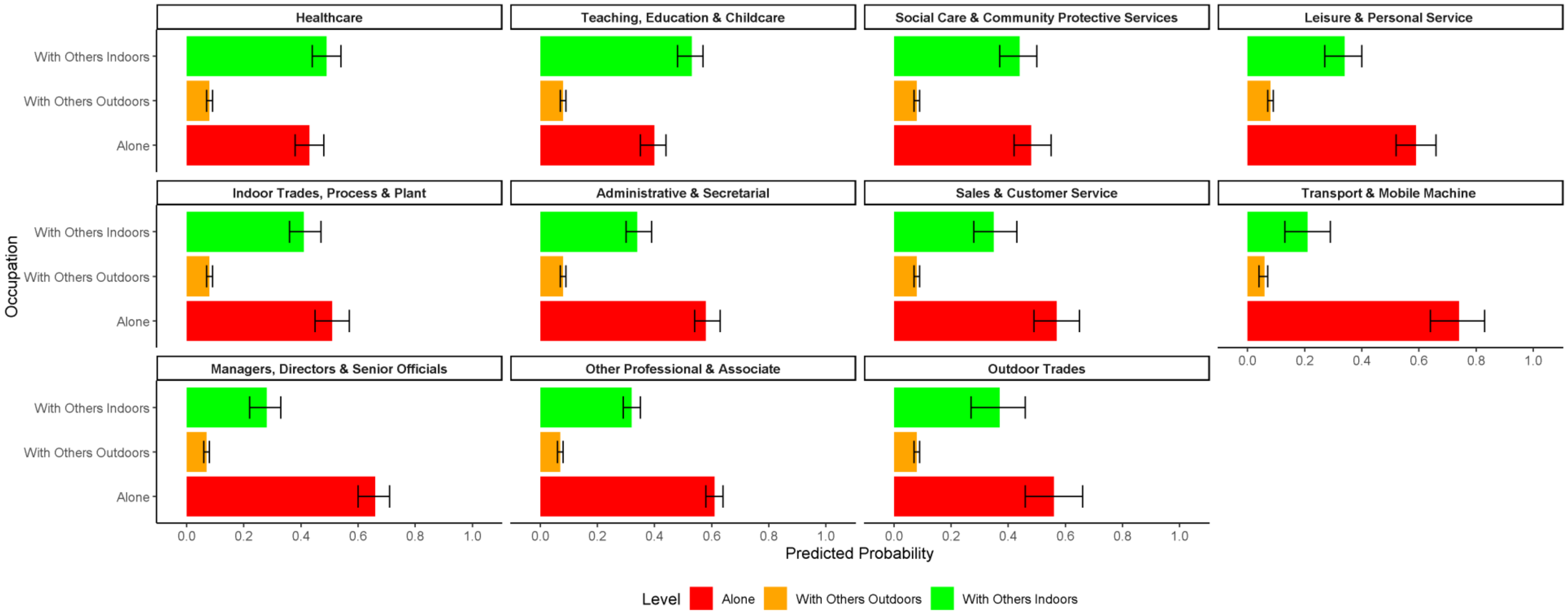

**Supplementary Figure 14. Frequency of Work-Related Social Gatherings: Predicted probabilities by occupation over time**

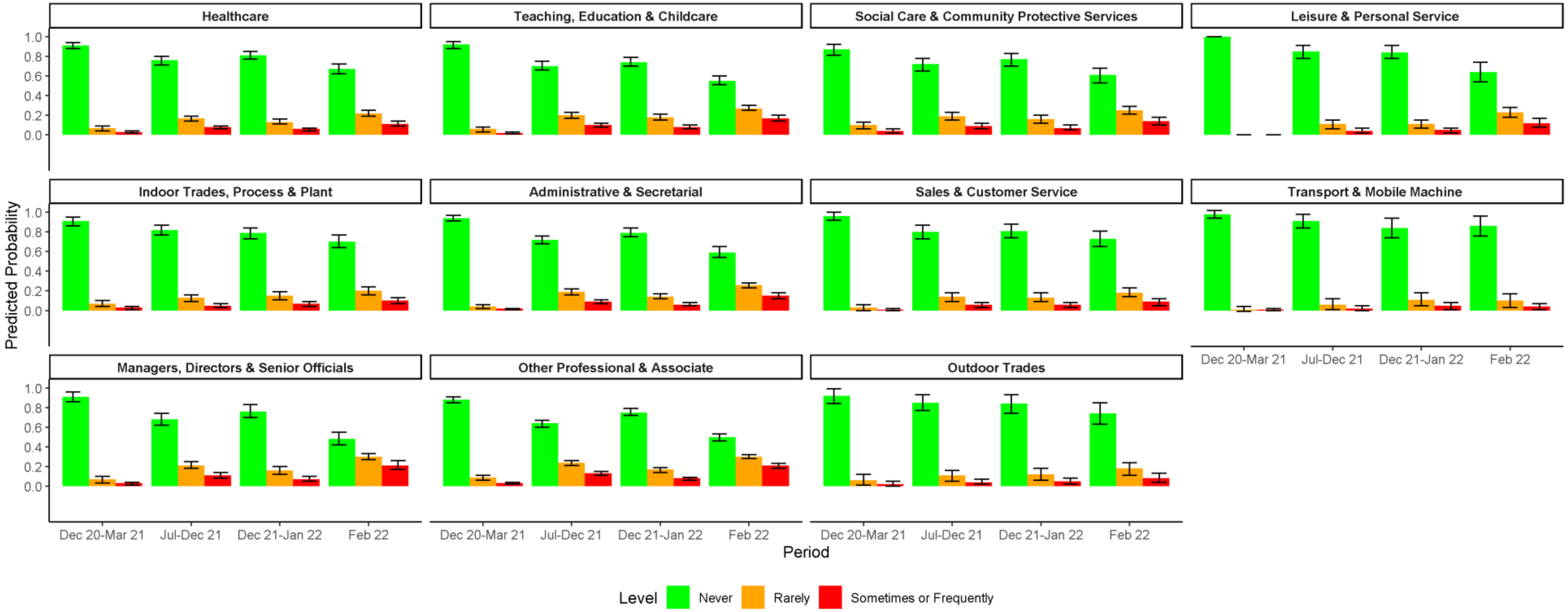

**Supplementary Figure 15. Workplace Provision of Lateral Flow Tests: Predicted probabilities by occupation**

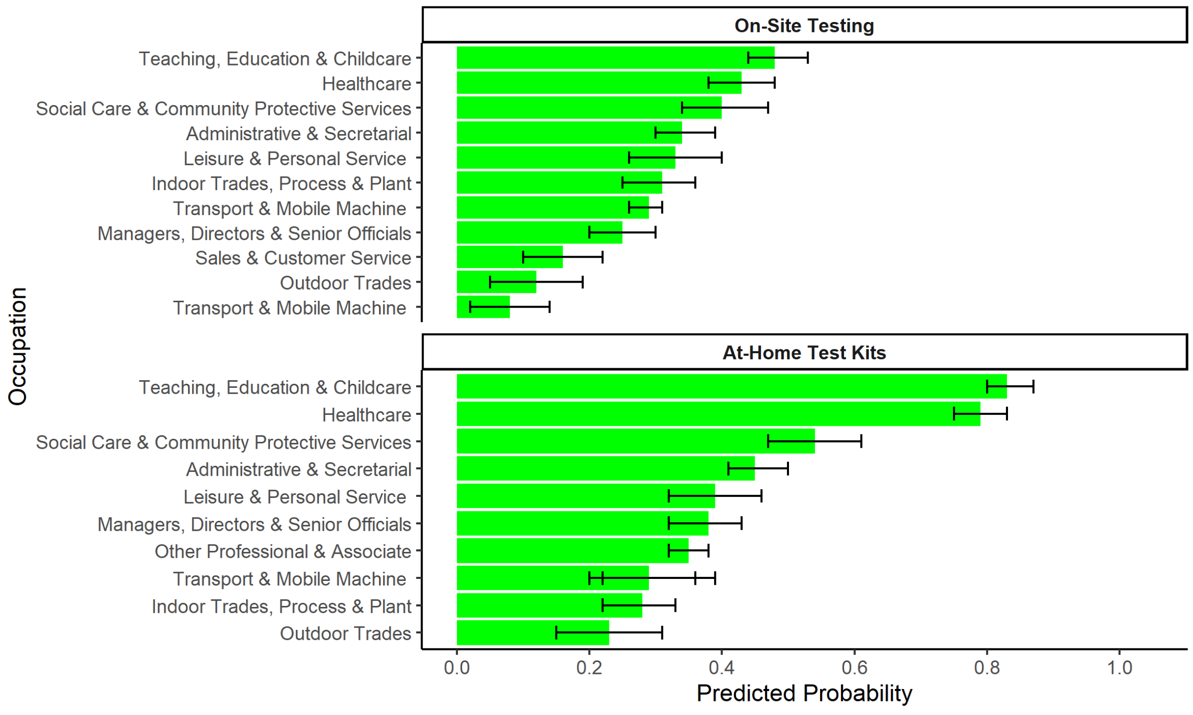
